# Anterior insula and mid-cingulate cortex differentially regulate anxiety and fear brain–body responses

**DOI:** 10.1101/2025.10.08.25337596

**Authors:** Jennifer Legon, Andrew Strohman, Yunrou Ni, Gabriel Isaac, Aditya Kapoor, Katheryn Painchaud, Sahib Khalsa, Wynn Legon

**Affiliations:** Fralin Biomedical Research Institute at Virginia Tech Carilion, Roanoke, VA, 24016, USA; School of Neuroscience, Virginia Polytechnic Institute and State University, Blacksburg, VA, 24061, USA; Virginia Tech Carilion School of Medicine, Roanoke, VA, 24016, USA; Graduate Program in Translational Biology, Medicine, and Health, Virginia Polytechnic Institute and State University, Roanoke, VA, 24016, USA; Center for Human Neuroscience Research, Fralin Biomedical Research Institute at Virginia Tech Carilion, Roanoke, VA, 24016, USA; Department of Psychiatry and Biobehavioral Sciences, Semel Institute for Neuroscience and Human Behavior, David Geffen School of Medicine, University of California at Los Angeles, Los Angeles, CA 90095, USA; Center for Health Behaviors Research, Fralin Biomedical Research Institute at Virginia Tech Carilion, Roanoke, VA, 24016, USA

## Abstract

Heightened reactivity to predictable and unpredictable threat is a hallmark of anxiety, yet causal evidence for the roles of deep interoceptive and salience-network hubs remains limited. Using low-intensity focused ultrasound (LIFU), we transiently modulated the ventral anterior insula (vAI) and anterior mid-cingulate cortex (aMCC) in healthy adults performing the No–Predictable–Unpredictable (NPU) threat task. We quantified anxiety-potentiated responses (APR) and fear-potentiated responses (FPR) along with brain–body markers including EMG startle responses, EEG event-related potentials, electrodermal responses and heart rate (HR). Mixed-effects models revealed a significant LIFU × Task interaction such that LIFU to the vAI reduced APR symptom reports and LIFU to the aMCC reduced FPR symptom reports. Physiological changes largely paralleled behavior: vAI modulation reduced APR-linked startle, EEG response, and HR, while aMCC effects were confined to FPR (EEG and HR reductions). Brain-Body-Behavior analysis revealed that LIFU to the vAI increased the coupling of both HR and EEG with subjective reports such that changes in HR and the EEG evoked potential predicted symptom ratings but only during the APR task. In addition, baseline HR/HRV correlated with anxiety across individuals but did not moderate these neuromodulation effects. These findings provide causal, site-specific evidence that the vAI preferentially supports anticipatory anxiety under uncertainty, whereas the aMCC contributes to brain–body mobilization for predictable threat. Targeted neuromodulation of the vAI increases brain-body-behavior coupling to reduce physiological response that translates to reduced symptoms. Neuromodulation targeting the vAI may enable precision interventions for anxiety-related disorders.

**Significance Statement:** Anxiety arises from dysregulated interactions between the brain and body, but causal evidence for the cortical hubs governing these loops has been lacking. Using low-intensity focused ultrasound (LIFU), we transiently modulated the ventral anterior insula (vAI) and anterior mid-cingulate cortex (aMCC) during a threat uncertainty task. vAI stimulation reduced anticipatory anxiety symptoms under uncertainty and strengthened heart rate and EEG coupling with subjective reports, consistent with aberrant interoceptive prediction processing. aMCC stimulation selectively dampened cue-evoked fear without enhancing brain–body alignment, consistent with a role in defensive mobilization. These dissociable effects across physiology, behavior, and report provide causal evidence for interoceptive–autonomic integration and highlight circuit-specific neuromodulation as a potential therapy for anxiety.

## Introduction

Anxiety is characterized by heightened sensitivity to threat and uncertainty, often manifesting as exaggerated defensive responses to both predictable and unpredictable aversive events. Converging evidence implicates the ventral anterior insula (vAI) and anterior mid-cingulate cortex (aMCC) as key hubs linking salient stimuli to emotional and bodily states through predictive processing and interoceptive integration (1–5). These regions are central nodes buried deep within the salience, action mode, and central autonomic networks (3,6–10) coordinating cognitive appraisal with autonomic and behavioral mobilization to respond to environmental demands, making them promising therapeutic targets for anxiety conditions.

Contemporary models suggest that dysfunction within the vAI and aMCC contributes to impaired interoceptive awareness, threat appraisal, and defensive responding under uncertainty (11,12). The vAI encodes the subjective intensity and aversiveness of internal states, integrating affective and autonomic signals to generate anticipatory anxiety (13), whereas the aMCC is more closely tied to action monitoring and preparation for responses to explicit threat cues (14,15). Both regions exhibit hyperactivity across anxiety disorders, consistent with increased vigilance and dysregulated autonomic reactivity (16,17). Yet causal evidence for their distinct contributions in humans remains limited.

Laboratory paradigms such as the No–Predictable–Unpredictable (NPU) threat task (18) provide an ideal testbed, reliably eliciting subjective anxiety, physiological arousal, and behavioral changes under conditions of uncertainty or predictable threat (19,20). Two derived indices capture dissociable processes: anticipatory anxiety-potentiated responses (APR) and fear-potentiated responses (FPR). APR, calculated as the difference between unpredictable and neutral no-cue conditions, indexes anticipatory anxiety under uncertainty. Elevated APR is associated with hypervigilance and intolerance of uncertainty (21), while reduced APR may signal resilience or successful regulation. FPR, defined as the difference between cued and un-cued predictable threat conditions, reflects cue-potentiated fear reactivity. Higher FPR suggests heightened reactivity to known threat cues, whereas lower FPR may reflect improved regulatory control (18,22).

In addition to behavioral anxiety ratings and reflexive measures like the startle reflex, neural and autonomic responses to predictable and unpredictable threat offer complementary insights into the mechanisms of anxiety and its modulation. Event-related potentials (ERPs) recorded via surface scalp electroencephalography (EEG) provide temporally precise markers of sensory and cognitive processing in response to threat cues (23,24). Both predictable and unpredictable threats enhance early and late ERP components which are thought to reflect increased attentional allocation, salience detection, and sustained emotional processing (24). Heightened ERP amplitudes to threat stimuli are consistently observed in clinical populations with anxiety and are sensitive to both acute stress and clinical state (25) and these ERPs have been linked to both the insula and the anterior cingulate cortex (26).

In addition to brain derived markers, cardiac and autonomic signals such as electrodermal responses can be sensitive measures for anxiety-related behaviors (27,28) and are strongly implicated in the development and maintenance of anxiety disorders (5,28–30). For example, individuals with high anxiety sensitivity and panic disorder show heightened cardiac interoception that increases the likelihood that benign physiological changes are misinterpreted as signs of threat (31). Furthermore, patients with anxiety disorders tend to have higher heart rates and lower heart rate variability (32–35) that may serve to signal anxious states. Both the insula and aMCC are critical hubs that receive and control cardiac and sympathetic signals (36–38) and the aberrant activity in these regions in patients with anxiety disorders may reflect dysfunctional brain-body integration. Moreover, existing pharmacotherapies are ineffective in improving these measures towards healthy levels(33,39,40).

A growing body of evidence suggests that non-invasive neuromodulation may be an effective means to reduce anxiety(41–44). Recent research has demonstrated that TMS can influence anxiety potentiated (APR) and fear potentiated (FPR) physiological and behavioral responses during the NPU task. Balderston et al. (2020)(22) showed that low-frequency (1 Hz) transcranial magnetic stimulation (TMS) to the parietal cortex reduced both facilitated predictable startle and anticipatory predictable startle, though subjective anxiety ratings remained unchanged. In two studies targeting the right dorsolateral prefrontal cortex, Teferi and colleagues found both continuous or intermittent theta burst stimulation facilitated anxiety-related startle responses(45,46). One reason TMS may not have produced the hypothesized decreases may be due to the inability to deliver energy directly to deeper brain regions like the insula and cingulate cortex implicated in anxiety-related behavioral and physiologic responses. Both TMS and transcranial electric stimulation (TES) suffer from a depth-focality tradeoff that limits energy to the surface of the cortex (47,48). New non-invasive methods to directly reduce activity in the vAI and/or aMCC during predictable or unpredictable threat may serve to differentially affect these physiological measures to reduce anxiety-related responses.

Low-intensity focused ultrasound (LIFU) is an emerging neuromodulation modality that uses mechanical energy to modulate neuronal activity(49). Unlike other non-invasive methods, LIFU can be precisely focused to deep brain regions with millimeter-scale spatial resolution(49). We have previously demonstrated that LIFU can target subregions of the insula and the aMCC to modulate pain-related behavior, in addition to brain event-related potentials, heart-rate variability and markers of brain-body interaction such as the heart-beat evoked potential (50–53). Building on these findings, we now seek to translate this approach to anxiety-related processes given that the insula and aMCC are key hubs of the salience and anxiety networks, integrating interoceptive, autonomic, and affective signals that shape threat anticipation and defensive responding. We hypothesized that LIFU targeting the vAI would preferentially reduce anxiety-potentiated responses (APR) due to its involvement in anticipation (54) and interoceptive predictive coding (1) whereas LIFU targeting the aMCC would preferentially reduce FPR due to its supporting role in preparation for defensive responses in the face of predictable threats (55,56). We targeted healthy individuals in this study as a first step towards investigating a role for vAI and aMCC neuromodulation in anxiety disorders.

## METHODS

### Participants

The Virginia Tech Institutional Review Board approved all experimental procedures (VT-IRB #23-192). N = 40 healthy participants (12M/28F, aged 26.2 ± 5.8 years); range (18–42), who met all inclusion/exclusion criteria and provided written informed consent to all aspects of the study and were financially compensated for participation. Inclusion criteria were females and males ages 18-45. Exclusion criteria were in accordance with contraindications to noninvasive neuromodulation as outlined by Rossi et al. (2009). Exclusions included contradictions for magnetic resonance imaging (MRI) and computed tomography (CT), a history of neurological disorders (e.g., Parkinson’s disease, epilepsy, multiple sclerosis, etc.), head injury resulting in loss of consciousness for >10 minutes, any current treatment or medical disorder with potential nervous system effects, pregnancy, history of alcohol or drug dependence, and/or history of or current cardiovascular disease.

### Overall Study Design

This was a single-blind, sham-controlled, cross-over design. Data were collected in four visits over 4 separate days with a minimum of 48 hours between each visit to mitigate any potential carry-over effects. Following informed consent, the first study visit comprised a structural brain MRI and CT scan for the purpose of low-intensity focused ultrasound (LIFU) targeting and acoustic modelling. The remaining three visits were formal testing sessions of LIFU to the right anterior insula (AI), right dorsal anterior cingulate cortex (aMCC), or sham, randomized between and within subjects (**Figure 1**).

**Figure 1.**
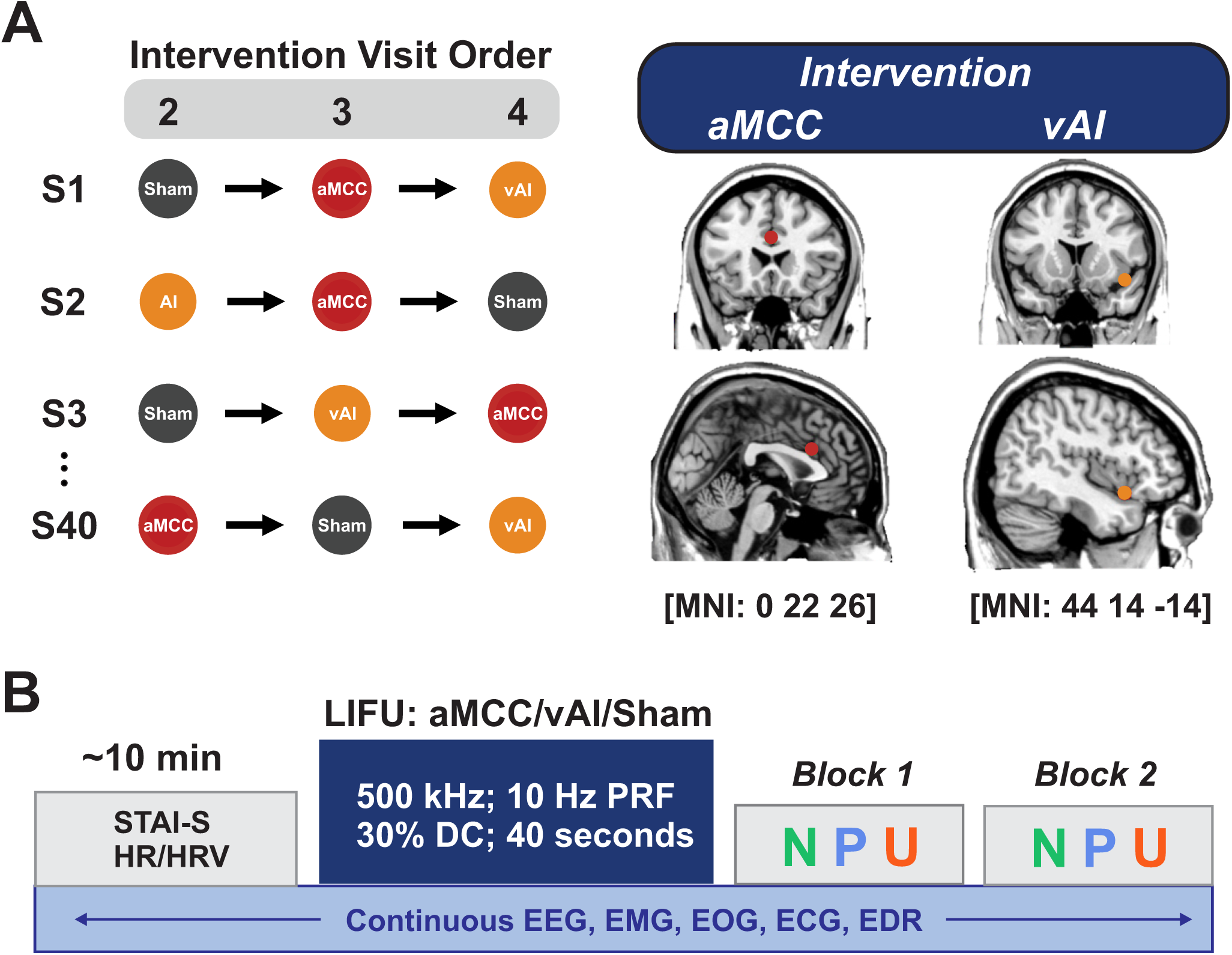
Overall Study Design. **A.** *Study visits and intervention conditions*. Participants (N = 40) underwent three randomized sessions with LIFU targeted to the right ventral anterior insula (vAI), right anterior mid-cingulate cortex (aMCC), or active sham. The schematic on the left shows counterbalanced intervention orders with and between participants. On the right, 5 mm ROIs are placed at MNI coordinate [0 22 26] for aMCC targeting and 5 mm ROI at MNI coordinate [44 14 -14] for vAI targeting. **B.** *Experimental timeline*. Each testing session (∼2 hr) queried state anxiety (STAI-S) and recorded baseline heart-rate (HR) and heart-rate variability (HRV). Low-intensity focused ultrasound (LIFU) was delivered for 40 seconds with parameters: 500 kHz, 10 Hz pulse repetition frequency (PRF) and 30% duty cycle. After LIFU, participants performed two blocks of the No-threat, Predictable-threat, and Unpredictable-threat (NPU) task. NPU block order (15 min each) were pseudorandomized across subjects, with brief breaks in between, and contained repeated trials with embedded acoustic startle probes and ratings of anxiety. Continuous physiological measures (EEG, EMG, EOG, ECG, EDR) and subjective ratings were collected throughout.

At each formal testing session (prior to LIFU), participants completed a review of symptoms (ROS) questionnaire, state anxiety inventory (STAI-S), and a daily activities questionnaire that queried sleep, caffeine intake, and physical activity on the day of testing. They were then seated in a comfortable chair with arm and neck support and connected to continuous electrooculogram (EOG), electromyogram (EMG), electroencephalogram (EEG), electrocardiogram (ECG), and electrodermal (EDR) monitoring (see below for details). After this, pain thresholding was conducted (see below) and a ∼10-minute baseline EOG, EMG, EEG, ECG, and EDR were collected while the participant rested in the chair. After this, startle habituation (18) was performed and after this participants underwent LIFU to either the AI, aMCC, or Sham for 40 seconds (57).

Roughly 5 minutes After LIFU subjects performed the No threat, Predictable Threat & Unpredictable Threat (NPU) task (18). Following the completion of the NPU task, 5 minutes of resting physiological data was additionally recorded. Subjects then completed a review of symptoms (ROS) questionnaire, state anxiety inventory (STAI-S) and a quality control questionnaire at least 30 minutes after LIFU application (see **Figure 1** for overall Experimental Design). Each session lasted approximately 2 hours. Detailed steps of each procedure are described below.

### MRI and CT Imaging

MRI data were acquired on a Siemens 3T Prisma scanner (Siemens Medical Solutions, Erlangen, Germany) at the Fralin Biomedical Research Institute’s Human Neuroimaging Laboratory. Anatomical scans were acquired using a T1-weighted MPRAGE sequence with a TR = 1400 ms, TI = 600 ms, TE = 2.66 ms, flip angle = 12°, voxel size = 0.5×0.5×1.0 mm, FoV read = 245 mm, FoV phase of 87.5%, 192 slices, ascending acquisition.

Computerized Tomography (CT) scans were also collected with a Kernel = Hr60 in the bone window, FoV = 250 mm, kilovolts (kV) = 120, rotation time = 1 second, delay = 2 seconds, pitch = 0.55, caudocranial image acquisition order, 1.0 mm image increments for a total of 121 images and scan time of 13.14 seconds.

### Physiological Preparation

#### Electroencephalogram (EEG)

Surface EEG was collected from the vertex channel (CZ) using a 10 mm silver-over-silver chloride cup electrode referenced to the right mastoid. Data were continuously acquired at a 1 kHz sampling rate using a DC amplifier (EEG100C & MP160, Biopac Systems, CA, USA) and AcqKnowledge 5.0 software (BioPac Systems, CA, USA). The scalp was prepared with a mild abrasive gel (Nuprep; Weaver and Company) and rubbing alcohol. Cup electrodes were filled with a conductive paste (Ten20 Conductive; Weaver and Company) and held in place with medical tape. Electrode impedances were verified (<50 kΩ) before recording. Data were stored on a PC for offline data analysis.

#### Electrocardiogram (ECG)

Two latex-free ECG electrodes (MedGel, MDSM611903) were attached symmetrically to the anterior surface of the bilateral forearms immediately distal to the antecubital fossa and grounded to the right elbow. ECG data were continuously collected and sampled at 1 kHz using a DC amplifier (ECG100D & MP160, Biopac Systems, CA, USA) and AcqKnowledge 5.0 software and stored on a PC for offline data analysis.

#### Electrodermal response (EDR)

Two silver-over silver chloride electrodes were applied to the distal second and third digits their right hand with isotonic sodium chloride cream (BIOPAC GEL101A) and secured with medical tape. EDR data were sampled at 1 kHz using a DC amplifier (EDA100D & MP160, Biopac Systems, CA, USA) with a constant voltage of 0.5 V and recorded using the AcqKnowledge 5.0 software and stored on a PC for offline data analysis.

#### Electrooculogram (EOG)

Two electrodes (Gereonics Inc., miniature silver/silver chloride 405 series) were placed underneath the right eye overlapping the orbicularis oculi muscle. Subjects were instructed to look forward and the first electrode (+) was positioned so that it was placed directly below the pupil. The second electrode (-) was placed 1mm laterally from the first electrode. The wells of the electrodes were filled with a small amount of conductive gel (Spectra 360, Parker Laboratories Inc NJ)

#### Aversive Stimulus Thresholding

The aversive stimulus for the NPU task was a heat pain stimulus. This thermal stimulus was delivered using a contact 3×3.2 × 2.4 mm peltier thermode (T03, QST.lab, Strasbourg, FR) and a cutaneous stimulator (TCS, QST.lab, Strasbourg, FR). At each formal testing visit (sessions 2-4) participants were familiarized with the stimulation device. The thermal probe was placed on one of two marked spots on the forearm within the medial antebrachial dermatome of the right forearm. The first spot was measured using a measuring tape and marked 10cm from the elbow. The second spot was marked 5cm from the first marked spot. The device delivered a series of 3 heat stimulations that lasted for 0.7 seconds each at a ramping speed of 30°C/s with an inter-stimulus interval (ISI) of 5 seconds. Participants were instructed to verbally rate their subjective pain response after each thermal stimulation on a scale from 0-10, with zero representing ‘no pain’ and 10 representing ‘worst pain imaginable.’ This process was repeated by gradually adjusting the temperature until subjects reported an average response of 7/10 from the 3 stimuli. This intensity was then used for the aversive stimulus in the NPU task. Aversive heat provides a controlled, ecologically valid nociceptive stimulus that engages cutaneous and deep nociceptive pathways. Compared to electrical shock, heat pain evokes a slower but sustained nociceptive signal allowing for graded calibration of perceived intensity across participants. Importantly, heat stimulation avoids the abrupt startle and strong autonomic artifact (e.g., EMG contamination, skin-conductance saturation) often induced by electrical shock, improving data quality for physiological recordings. Prior work shows that both modalities elicit comparable anticipatory anxiety and defensive responses when matched for subjective intensity, supporting the validity of heat as an aversive stimulus in threat-of-pain paradigms (58–60).

#### Startle habituation

Prior to LIFU administration, participants were habituated to the startle stimulus as recommended(18). Startle stimuli were square-wave white noise at 103dB for a 40ms duration delivered through over-the-ear headphones. 12 total stimuli were delivered with an ISI of 5 seconds.

#### No shock, Predictable Shock, Unpredictable Shock (NPU) threat task

The NPU task was conducted according to Schmitz & Grillon (18). The threat task consisted of three conditions, one in which no aversive stimulus was delivered (N), and two conditions where the aversive stimulus was delivered while a cue was present (predictable (P)) or at any time (unpredictable (U)). Prior to the task, subjects were given instructions and familiarized with cues and their corresponding conditions. Each condition (N, P, U) had a designated colorful cue. During each condition, cues appeared on a screen for 8 seconds with an ISI of 16 seconds. In addition to the cue on screen, instructions at the top of the screen informed participants what the condition was. In the ‘N’ condition, no aversive stimuli were delivered to the subject regardless of whether the cue was present or not. In the ‘P’ condition, the aversive stimuli were delivered only when the cue appeared on the screen. In the ‘U’ condition, the aversive stimulus could be delivered at any time regardless of the cue.

The experiment consisted of two separate task blocks. Task block 1 was ordered P-N-U-N-U-N-P and task block 2 was ordered U-N-P-N-P-N-U. There was a startle habituation period prior to each task block. Each task block lasted approximately 15 minutes. The order of task blocks was randomized between subjects. There was a 5-minute break between task blocks. For each letter within a task block, there were three trials. At the end of the third trial, participants were prompted to rate their perceived anxiety symptoms on a scale of 0-10, where 0 indicated ‘not anxious’ and 10 indicated ‘extremely anxious’ for both when the cue was present or not present (**Figure 1**). Within any given letter trial, there were two randomly presented startle probes with a minimum ISI of 10 seconds. For any given letter trial, there were two startle probes. For any given task block, there were 42 (7×3×2) startle probes.

### Assessments

Prior to testing all participants filled out the State and Trait Anxiety Inventory (STAI-T)(61), the Beck Depression Inventory (BDI) (62), the Beck Anxiety Inventory (BAI) (63), the International Physical Activity Questionnaire (IPAQ) (64), the Medical Outcomes Survey Short Form-8 (SF-8), the sleep survey/sleep scale from the Medical Outcomes Study, the Patient Health Questionnaire (PHQ2), the Generalized Anxiety Disorder 2-item (GAD2), the Tobacco, Alcohol, Prescription medications & other Substances Tool (TAPS), and the Perceived Stress Scale (PSS).

#### STAI State

The State Trait Anxiety Inventory (STAI)(61) was used to assess trait and state anxiety through a series of questions querying participants on feelings of anxiety and their perceived severity. Participants rated the intensity of their feelings using a 4-point Likert scale: not at all, somewhat, moderately so, very much so. The state version of this questionnaire was collected at the beginning of each visit (sessions 2–4) and a minimum of 30 minutes after the LIFU or sham application.

#### Review of Symptoms

A review of symptoms questionnaire(65) queried participants about the presence of various symptoms and their severity (absent/mild/moderate/severe) scored on a scale of 0–3. This questionnaire was collected at the beginning of each visit (sessions 2–4) and a minimum of 30 minutes after the LIFU or sham application to indicate any changes of symptoms from the intervention. The symptom questionnaire is provided in Supplementary data.

### LIFU Transducer

For the vAI we used a Sonic Concepts H-281 single-element 500 kHz transducer with an active diameter of 45.0 mm and a geometric focus of 45.0 mm. The focal depth from the exit plane was 38.0 mm. The transducer also had a solid water coupling over the radiating surface to the exit plane. For the aMCC we used a Sonic Concepts H-104 single-element 500kHz transducer with an active diameter of 64mm and a geometric focus of 63.2mm. The focal depth from the exit plane was 52mm.

#### LIFU waveform

LIFU waveforms were generated using a two-channel, 2-MHz function generator (BK 4078B Precision Instruments). Channel 1 was used to gate channel 2 which was a 500 kHz sine wave. Channel 1 was a tapered 5Vp-p square wave burst of 10 Hz (N = 400) with a pulse duration of 30 milliseconds and a pulse repetition interval of 100ms. This resulted in a 40-sec total LIFU application time to each brain region with a duty cycle of 30% (66,67). The output of channel 2 was sent through a 100-W linear RF amplifier (E&I 2100L; Electronics & Innovation) before being sent to the LIFU transducer. The peak negative pressure of the waveform outside the head was ∼900 kPa or 27 W/cm^2^ spatial peak pulse average intensity (I_SPPA_) for all participants.

### LIFU targeting

The transducer was coupled to the head using conventional ultrasound gel and custom mineral oil/polymer coupling pucks (68). These pucks have negligible attenuation at 500 kHz and can be made with varying stand-off heights that allow for precise axial (depth) targeting based on individual target depths.

Each participant’s right vAI and aMCC targets were based on meta-analytic evidence of the convergence of induced and pathological anxiety during unpredictable shock (69) and automated meta-analysis based on the search term ‘anxiety’ (70). The right vAI target was Montreal Neurologic Institute (MNI) coordinate [44 14 -14] identified with the aid of an insular atlas [46] while the right aMCC target was MNI coordinate [0 22 26] (**Figure 2**). Depth to each target was measured from the scalp. An appropriate coupling puck was made so that the focal spot of the transducer was overlaid on the target. Placement of the transducer on the scalp was aided using a neuronavigation system (BrainSight, Rogue Research, Montreal, QUE, CAN). LIFU or Sham was only delivered if placement error was < 3 mm.

**Figure 2.**
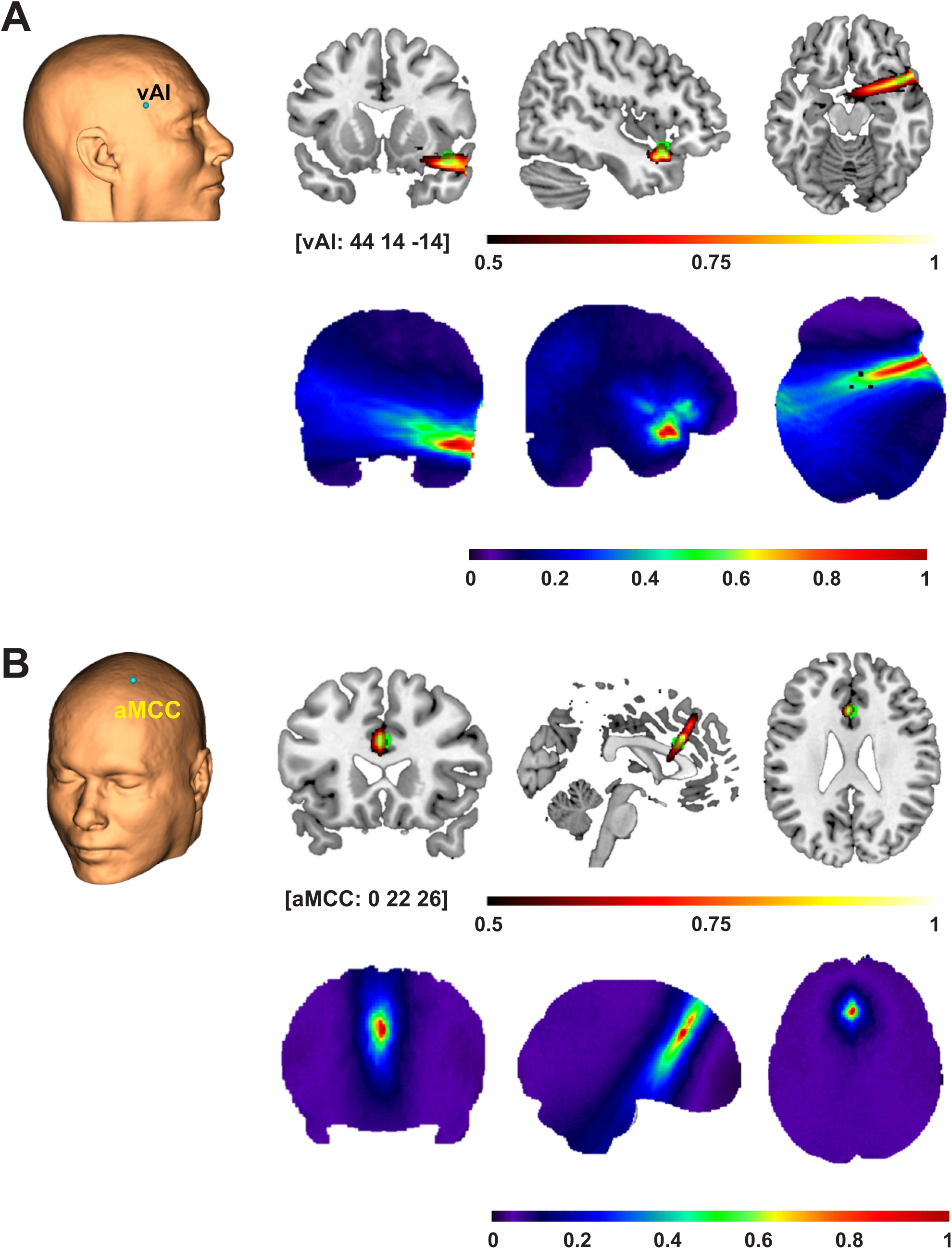
LIFU targeting the anterior insula and anterior mid-cingulate cortex (aMCC) **A.** Scalp rendering showing placement of the center of the transducer (blue dot) to target the right ventral anterior insula (vAI) [MNI: 44 14 -14] from one participant. (Top Right) Group (N = 40) normalized acoustic pressure (warm colors) showing top 50% energy distribution in MNI space. A 5mm ROI sphere is also shown centered on coordinate [44 14 -14] to show targeting accuracy to scale. Scale bar is normalized pressure. (Bottom Right) Group (N = 40) normalized pressure map showing full pressure distribution across the group at scale with above brain images. Scale bar is normalized pressure. **B.** Scalp rendering showing placement of the center of the transducer (blue dot) to target the right anterior mid-cingulate cortex (aMCC) [MNI: 0 22 26] from one participant. (Top Right) Group (N = 40) normalized acoustic pressure (warm colors) showing top 50% energy distribution in MNI space. A 5mm ROI sphere is also shown centered on coordinate [0 22 26] to show targeting accuracy to scale. Scale bar is normalized pressure. (Bottom Right) Group (N = 40) normalized pressure map showing full pressure distribution across the group at scale with above brain images. Scale bar is normalized pressure.

### Model Heating

Thermal models were performed using an adapted modified mixed-domain method (mSOUND)(71) with a 3-layer skull model (cortical-trabecular-cortical) from Benchmark 6 (72) to solve for the temperature fields based on the acoustic field and the bioheat equation using k-wave diffusion (73). This model assumes that 100% of the attenuation is absorption and thus converted to heat and, as such, is a conservative estimate of heating. We performed the thermal simulation for 30 ms on and 70 ms off (30% duty cycle) for 40 seconds using 1 MPa input pressure.

### Sham condition

The Sham condition involved an active sham, where all procedures were identical to LIFU visits (including the use of neuronavigation, the selection of the proper gel puck, and the application of continuous auditory masking), except a high impedance material was inserted into the gel that attenuated the ultrasound prior to reaching the skull. For the Sham visit, the transducer was placed at one of the active sites, with the site randomized between participants. An auditory masking questionnaire was used after each session to query on successful masking (see next section).

### Acoustic masking

In some cases, a single-element transducer can produce an audible auditory artifact likely as the result of the pulse repetition frequency. To address this potential confound (74), strict acoustic masking was performed. Acoustic masking was delivered through disposable earbuds that were plugged into a tablet. A combination of sounds from a white noise app were then randomly mixed creating a multitone that has previously been demonstrated to effectively mask ultrasound artifact (75). Participants were told to set the volume to a comfortable level that removed ambient sounds. This intensity range was on average 70 – 75 dB. Auditory masking was confirmed by speaking to the participant outside of their visual field. Auditory masking noise was played continuously throughout the testing session. 30 minutes after formal testing, participants were queried on auditory masking. Questions included “I could hear the LIFU stimulation”, “I could feel the LIFU stimulation”, and “I believe I experienced LIFU stimulation.” Participants were asked to respond to each question using a 7-point Likert scale (0–6) with points corresponding to Strongly Disagree / Disagree/ Somewhat Disagree / Neutral / Somewhat Agree / Agree / Strongly Agree.

### Preprocessing & Analysis

#### Anxiety Response

Anxiety symptoms were quantified for cue and ITI (inter-trial interval) periods for each of the N, P, U trials for each participant. We followed the methods of Balderston et al. (2020)(22) and calculated the APR (Anxiety-potentiated response) and FPR (Fear-potentiated response). For APR, we subtracted the anxiety rating during the neutral ITI from the anxiety rating during the unpredictable ITI. For FPR, we subtracted the anxiety rating during the predictable ITI from the anxiety rating during the predictable cue. We fit linear mixed-effects models with fixed effects of LIFU (vAI, aMCC, Sham), TASK (APR, FPR) and their interaction and random intercepts and TASK slopes by subject: VAS ∼ LIFU * TASK + (1 + TASK | Subject).

Planned contrasts included examining APR and FPR for vAI and aMCC versus Sham. All p-values were corrected using the Benjamini-Hochberg (BH) method. The same model was used for each of the univariate physiological outcome measures below and corrected for false discovery rate using BH.

#### Electromyographic (EMG) Startle Response

The EMG data stream was band-pass filtered from 30 to 250 Hz using a 4^th^ order Butterworth filter and additionally filtered using a Butterworth 2^nd^ order notch filter from 59 – 61 Hz using the `filtfilt` function in Matlab® (R2024a). The data was then epoched around the startle events [-4 4] seconds. The area-under-the-curve (AUC) was then calculated from the time-window [0.05 0.25] seconds from the rectified EMG signal. These data were parsed into respective bins (N cue, N ITI, P cue, P ITI, U cue, UITI) depending on their timing relative to the cue for each N, P, U condition. This resulted in 18, 18, 12, 12, 12 and 12 events for each of the respective conditions. The mean for each of these conditions was calculated for each participant and used for subsequent statistical testing. APR and FPR were calculated from these means.

#### Electrodermal Response

The electrodermal data stream was band-pass filtered from 0.5 to 5 Hz using a 2^nd^ order Butterworth filter and the Matlab function `filtfilt`. The data was then epoched around both the startle events and the aversive events from [-5 10] seconds. The area-under-the-curve (AUC) was then calculated from the time-window [1 4.5] seconds from the rectified EDR signal. These data were then parsed into the respective N, P and U ITI and Cue bins resulting in the same number of events per bin as for the EMG data. The mean for each of these conditions was calculated for each participant and used for subsequent statistical testing. APR and FPR were calculated from these means and these data were used for statistical testing.

#### Heart-rate Response

Heart-rate data was taken from the Biopac® ongoing HR channel in the AcqKnowledge 5.0 software that provides a continuous, beat-to-beat representation of heart-rate in beats per minute. The ongoing HR is calculated using the interval between successive R-waves of the raw electrocardiogram (ECG) trace. These data were inspected for artifact and any aberrant data points (> or < 3 SD of the mean data stream) were removed and missing data interpolated using nearest-neighbor and the `interp` function in Matlab. The data were then epoched around startle events from [-5 5] seconds. Similar to above physiological data, these epochs were parsed into respective N, P and U ITI and Cue bins and the mean of the events was calculated for each participant for each bin. APR and FPR were calculated from these means for each participant and these data were used for statistical testing.

#### Electroencephalography (EEG) Response

EEG recordings were bandpass filtered from 0.1 to 55 Hz using a 3^rd^ order Butterworth filter and the Matlab function `filtfilt.` The data were then epoched around both the startle and aversive events from -2 to 5 seconds. For the startle events, the P and U had 24 events and the N condition had 36 events. All EEG data was inspected for blink and muscle artifact and any removed accordingly. Peak-to-peak analysis was performed using automated scripts written in Matlab to look for maxima and minima with a [0.1 0.5] time window. These time windows were chosen based upon the data and standardized across participants and conditions. For all ERP analysis the data was low-pass filtered at 10 Hz using a 3^rd^ order Butterworth filter and the function `filtfilt` in Matlab. All data was baseline normalized using [-1 0] baseline window. The response from each event for each condition was manually inspected and averaged so that each participant had a single averaged time series EEG that was used for quantification and statistical testing.

### Brain-Body-Behavior Coupling

To test whether within-session fluctuations in physiological measures covaried with subjective anxiety and whether this coupling depended on task (APR vs FPR) and LIFU target (AI vs aMCC); we fit mixed-effects models to sham-difference scores (AI–Sham; aMCC-Sham) and included a within-person physiology term (PhysWithin). Unlike the primary outcome models (which assess level differences vs Sham), these coupling models estimate slopes linking changes in physiology relative to sham to concurrent changes in symptom ratings relative to sham, and tested whether those slopes differ by TASK and LIFU using the model: VAS ∼ LIFU x TASK x PhysWithin + (1 + TASK + PhysWithin + TASK:PhysWithin | Subject). Thus, these coupling analyses asked whether within-person deviations in physiology predicted corresponding deviations in subjective anxiety VAS relative to sham. That is, on occasions when a participant’s physiology was higher or lower than their own typical level for a given task, did their APR or FPR VAS also shift up or down? To compute these relationships, physiological predictors were centered within each participant and task and then standardized, so that slopes reflected whether a participant’s physiology was higher or lower than their own typical level in that condition. The models also incorporated random slopes for physiology and task-by-physiology terms, allowing coupling effects to vary across individuals rather than assuming a single fixed slope. Consequently, a significant effect in this framework would indicate that LIFU neuromodulation altered the strength or direction of brain–body–behavior coupling relative to Sham, rather than simply shifting the overall amplitude of the physiological response. We controlled the false discovery rate using Benjamini–Hochberg across modalities for omnibus terms and within modality for planned contrasts.

### Baseline physiology

#### Baseline HR and HRV

Baseline HR data was taken from a baseline window preceding formal testing of roughly 10 minutes where participants sat quietly. HR data was cleaned using a robust mediant absolute-deviation filter. Samples were flagged as outliers when |x_i_ – *m | > T* x MAD with *T = 3* and MAD = median (|x – m|). Flagged points were imputed with the value of the nearest non-outlier sample along the time index using nearest neighbor interpolation preserving series length and avoiding local smoothing. To test whether baseline physiology influenced LIFU effects, we fit linear mixed-effects models for APR and FPR with a subject random intercept and fixed effects of LIFU (AI, aMCC, Sham), plus both between-person (subject mean across sessions) and within-person (session deviation from that subject’s mean) components of each moderator (HR, RMSSD, SDNN). The model was: β_0_ + LIFU_j_ + M_b,i_ x LIFU_j_ + M_w,i,j_ x LIFU_j_ + u_i_ + ε_ij_where β_0_ is the fixed intercept; _i_ is the subject index; _j_ is the session/condition index (three LIFU levels: AI, aMCC, Sham); M_b,i_ is the between-subject moderator and M_w,i,j_ is the within person moderator; u_i_ is the subject-level random intercept and ε_ij_ is the residual error.

### Acoustic Modelling

Computational models were developed using individual subject MR and CT images to evaluate the wave propagation of LIFU across the skull and the resultant intracranial acoustic pressure maps. Simulations were performed using the k-Wave MATLAB toolbox(73), which used a pseudospectral time domain method to solve discretized wave equations on a spatial grid. CT images were used to construct the acoustic model of the skull, while MR images were used to target LIFU at either the AI or aMCC target, based on individual brain anatomy. Details of the modeling parameters can be found in Legon et al. (2018)(67). CT and MR images were first co-registered and then resampled for acoustic simulations at a finer resolution and the acoustic parameters for simulation were calculated from the CT images. The skull was extracted manually using a threshold intensity value and the intracranial space was assumed to be homogenous as ultrasound reflections between soft tissues are small(76). Acoustic parameters were calculated from CT data assuming a linear relationship between skull porosity and the acoustic parameters(77,78). The computational model of the ultrasound transducer used in simulations was constructed to recreate empirical acoustic pressure maps of focused ultrasound transmitted in the acoustic test tank similar to previous work(76).

## RESULTS

### Ultrasound beam characteristics

The ultrasound beam as measured in free water had a lateral full-width at half maximum (FWHM) resolution in the X plane of 3.3 mm and in the Y plane of 3.4 mm at the Z maximum. FWHM represents the area that received > 50% of peak energy. The axial FWHM was 23 mm ranging from - 10 mm to + 13 mm from the point of maximum pressure (38 mm from the exit plane) conferring an effective axial FWHM of 28 – 51 mm. The constructed model waveform used for all acoustic simulations was in good agreement with these empirical measurements validating its use in the models as has been demonstrated previously for both insula and aMCC targets (50,51,53).

### Ultrasound heating

The maximum heating was 37.8 D in trabecular bone (Δ 0.8 D) and 37.4 D (Δ 0.4 D) in the brain.

### Targeting Depths & Accuracy

The average depth of the AI target for males and females was: 39.0 ± 2.8 mm and 36.4 ± 3.4 mm respectively. The average depth of the aMCC target for males and females was: 49.9 ± 3.2 mm and 46.7.4 ± 2.5 mm (see Supplementary Table 1).

The mean ± SD of the transducer placement on the head from the prescribed spot for right AI and aMCC was: 0.64 ± 0.63 mm and 0.87 ± 0.73 mm respectively. A paired t-test revealed no significant differences t(39) = 1.51, p = 0.14. (See Supplementary Figures 1 & 2).

### Auditory Masking

We converted participant responses into numerical values 0 – 6 where 0 = strongly disagree, 1 = disagree, 2 = somewhat disagree, 3 = neutral, 4 = somewhat agree, 5 = agree and 6 = strongly agree. The mean ± SD and median (in brackets) for the query “I could hear LIFU” for AI, aMCC and Sham was: 1.4 ± 1.9 (0); 1.7 ± 2.0 (1) and 1.3 ± 1.8. The mean ± SD and median (in brackets) for the query “I could feel LIFU” for AI, aMCC and Sham was: 1.4 ± 1.7 (1); 1.5 ± 1.6 (1) and 1.2 ± 1.6 (1). The mean ± SD and median (in brackets) for the query “I believe I experienced LIFU” for AI, aMCC and Sham was: 3.5 ± 1.6 (3); 3.4 ± 1.3 (3.5) and 3.1 ± 1.5 (3). No statistical differences were found for any of the queries (all p-adjusted > 0.34). See Supplementary Figure 3.

### Trait Anxiety (STAI-T)

Participants filled out the STAI-T(61) on the first visit. The mean ± SD was 40.3 ± 11.3 with a range of 25 – 68 (see **Figure 3A**).

**Figure 3.**
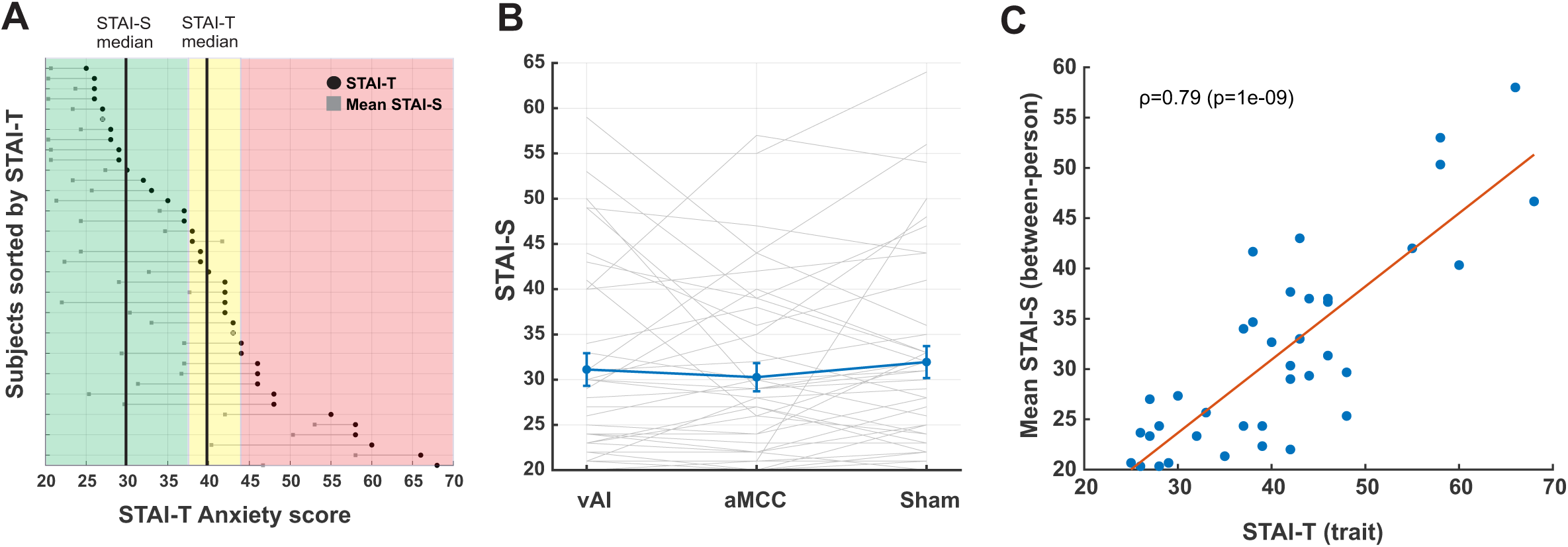
State and trait anxiety. **A.** Cleveland dot graph showing individual participants STAI-T (black dot) and mean STAI-S across sessions (grey square) ordered from highest to lowest STAI-T. The green, yellow, and red shaded regions illustrate suggested cut-offs for low, moderate and high STAI-S or STAI-T scores respectively. The vertical black lines denote the median for each of STAI-S and STAI-T of this cohort. **B.** Within-subject STAI-S scores across LIFU conditions (vAI, ACC, Sham). Grey lines denote individual subject responses. Blue line is group mean ± SEM. No differences in STAI-S was found across LIFU conditions (p = 0.74). **C.** A strong positive association was observed between STAI-T and mean STAI-S (ρ = 0.79, p = 1e–09), indicating that participants with higher trait anxiety consistently reported higher state anxiety across sessions.

### State Anxiety (STAI-S)

Participants filled out the STAI-S 30 minutes before and after each formal LIFU NPU session. The mean ± SD of all participants prior to LIFU for AI, aMCC and Sham was: 31.1 ± 11.4, 30.3 ± 9.8 and 31.9 ± 11.1. The mean ± SD of all participants after LIFU for AI, aMCC and Sham was 37.9 ± 9.4, 36.5 ± 9.1 and 37.6 ± 8.9. The respective mean ± SD change scores were: 6.7 ± 7.4, 6.2 ± 5.9 and 5.6 ± 8.8. See **Figure 3A** for Cleveland dot graph showing individual subject STAI-T and mean STAI-S across sessions. To examine if LIFU or the NPU task affected state anxiety we performed a one-way repeated measures analysis of variance (ANOVA) on the change scores. The one-way repeated measures ANOVA revealed no main effect: F(2,78) = 0.30, p = 0.74 (**Figure 3B**). There was a strong positive relationship (ρ = 0.79, p = 1e-09) between STAI-T and mean STAI-S (**Figure 3C**).

### Primary behavioral outcomes (APR & FPR)

We quantified fear and anxiety from the subjective anxiety ratings using APR and FPR where APR = Unpredictable No Cue – No Cue and FPR = Predictable Cue – Predictable No Cue as previously described (22). The mean ± SEM of APR for each of vAI, aMCC and Sham conditions was: 1.59 ± 0.22, 2.75 ± 0.27 and 2.87 ± 0.33. For FPR, the mean ± SEM for each of vAI, aMCC and Sham conditions was: 1.91 ± 0.21; 1.71 ± 0.21 and 2.74 ± 0.30 (see **Figure 4A**). We fit linear mixed-effects models with fixed effects of LIFU (AI, aMCC, Sham), TASK (APR, FPR) and their interaction, and random intercepts and TASK slopes by subject: VAS ∼ LIFU * TASK + (1 + TASK | Subject). The raw symptom ratings for N, P and U conditions separated into Cue and ITI conditions is presented in **Figure 4A** and Supplementary Table 2. The model showed a significant main effect of LIFU (F(2,200)=17.00, p=1.5×10⁻LJ) and a significant LIFU×TASK interaction (F(2,200)=8.22, p=3.7×10⁻LJ), with no TASK main effect (p=0.61).

**Figure 4.**
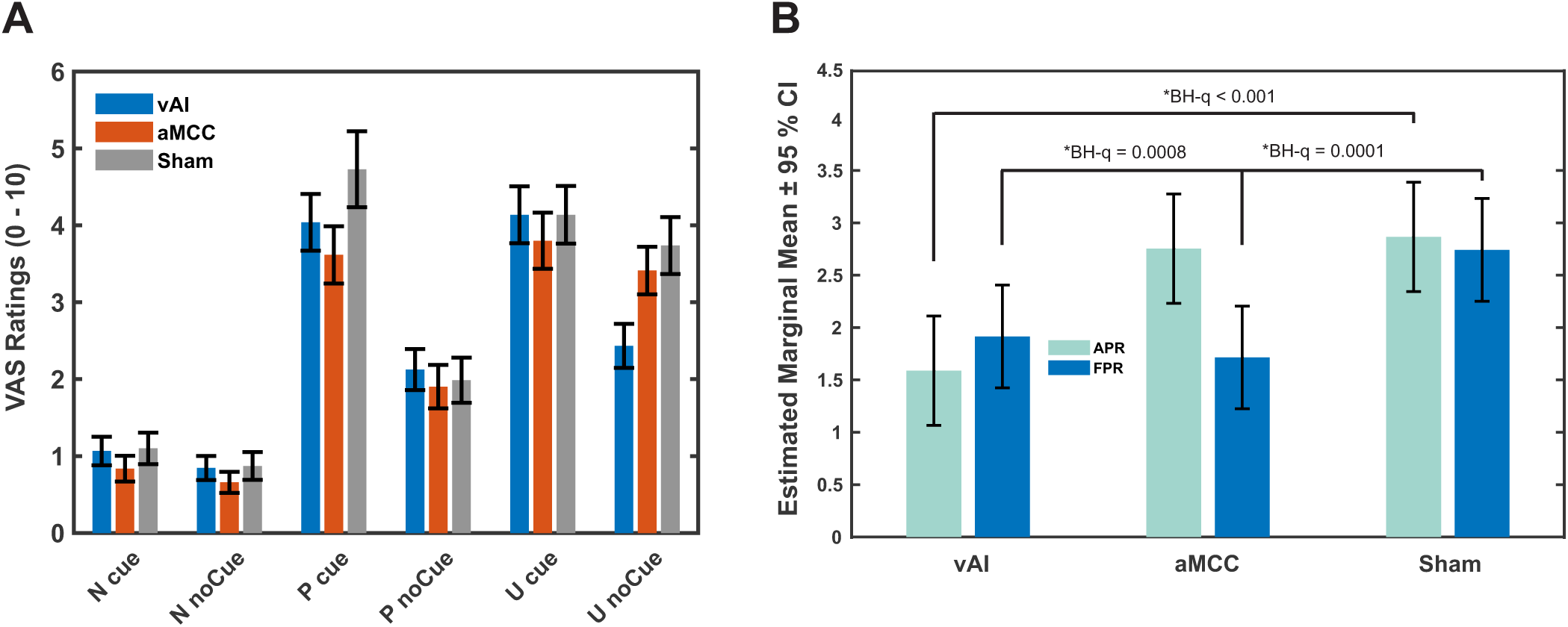
Behavioral effects of LIFU on anxiety-potentiated (APR) and fear-potentiated (FPR) responding. **A.** Raw VAS ratings for each N, P and U condition for each LIFU condition. LIFU to the ventral anterior insula (vAI) is in blue; LIFU to the anterior middle cingulate cortex (aMCC) is in red; Sham LIFU is in grey. **B.** Bars show estimated marginal means (EMMs) ± 95% confidence intervals from the linear mixed-effects model (VAS ∼ LIFU × TASK + (1+TASK|Subject)). APR is in light blue and FPR is dark blue. APR = U noCue – N noCue; FPR = P Cue – P noCue. The model estimates indicate how LIFU (vAI, aMCC) compares to Sham for each task condition, with error bars reflecting population-level uncertainty. * BH-q denotes significant Benjamini-Hochberg adjusted p-values.

Population-level marginal means indicated that vAI LIFU reduced both APR and FPR relative to Sham (APR: Δ= −1.28 ± 0.24, t(234)=−5.27, q<0.001; FPR: Δ=−0.83 ± 0.24, t(234)=−3.40, BH-q=0.0008), whereas aMCC LIFU selectively reduced FPR (Δ= −1.03 ± 0.24, t(234)= −4.23, BH-q=0.0001) but not APR (Δ= −0.11 ± 0.24, BH-q=0.64). Thus, LIFU to the ventral anterior insula preferentially attenuated APR, while LIFU to aMCC preferentially attenuated FPR symptom ratings (**Figure 4B**).

### Primary physiological outcomes

#### EMG Startle Response

The mean ± SEM EMG area under the curve (AUC) of the EMG startle response for APR for each of vAI, aMCC and Sham conditions was: 4.5 ± 5.2, 26.9 ± 5.3 and 20.8 ± 4.2 For FPR, the mean ± SEM for each of vAI, aMCC and Sham conditions was: 19.4 ± 5.2; 12.4 ± 5.8 and 20.1 ± 4.9. All values are in arbitrary units (AU). Raw mean ± SEM are in **Supplementary Table 3 & Supplementary** Figure 4. We fit linear mixed-effects models with fixed effects of LIFU (vAI, aMCC, Sham), TASK (APR, FPR) and their interaction, and random intercepts and TASK slopes by subject: EMG ∼ LIFU * TASK + (1 + TASK | Subject). The model showed a significant main effect of LIFU (F(2,200) = 7.35, p = 0.0008), no main effect of TASK (F(1,198.5) = 0.01, p = 0.906) and a significant LIFU × TASK interaction (F(2,200) = 5.91, p = 0.0032). Planned contrasts indicated that during APR, EMG responses were significantly reduced following AI LIFU compared to Sham (est = –16.40, SE = 6.08, t(234) = –2.70, p = 0.0075, q = 0.0149). aMCC LIFU did not differ significantly from Sham during APR (est = 6.13, p = 0.314). In the FPR condition, neither vAI nor aMCC stimulation differed significantly from Sham (AI vs Sham: est = –0.71, p = 0.907; aMCC vs Sham: est = –7.71, p = 0.206).

Estimated marginal means confirmed this pattern: Sham responses were similar across APR (M = 20.87, 95% CI [11.16, 30.57]) and FPR (M = 20.15, 95% CI [9.99, 30.31]). AI stimulation markedly reduced EMG during APR (M = 4.46, 95% CI [–5.25, 14.17]) but not FPR (M = 19.43, 95% CI [9.27, 29.60]). Conversely, aMCC stimulation increased APR responses (M = 27.00, 95% CI [17.29, 36.70]) and decreased FPR responses (M = 12.44, 95% CI [2.27, 22.60]), though neither effect reached significance. Together, these results indicate that AI LIFU selectively suppressed APR responses, while aMCC LIFU did not significantly modulate EMG relative to Sham (**Figure 5A**).

**Figure 5.**
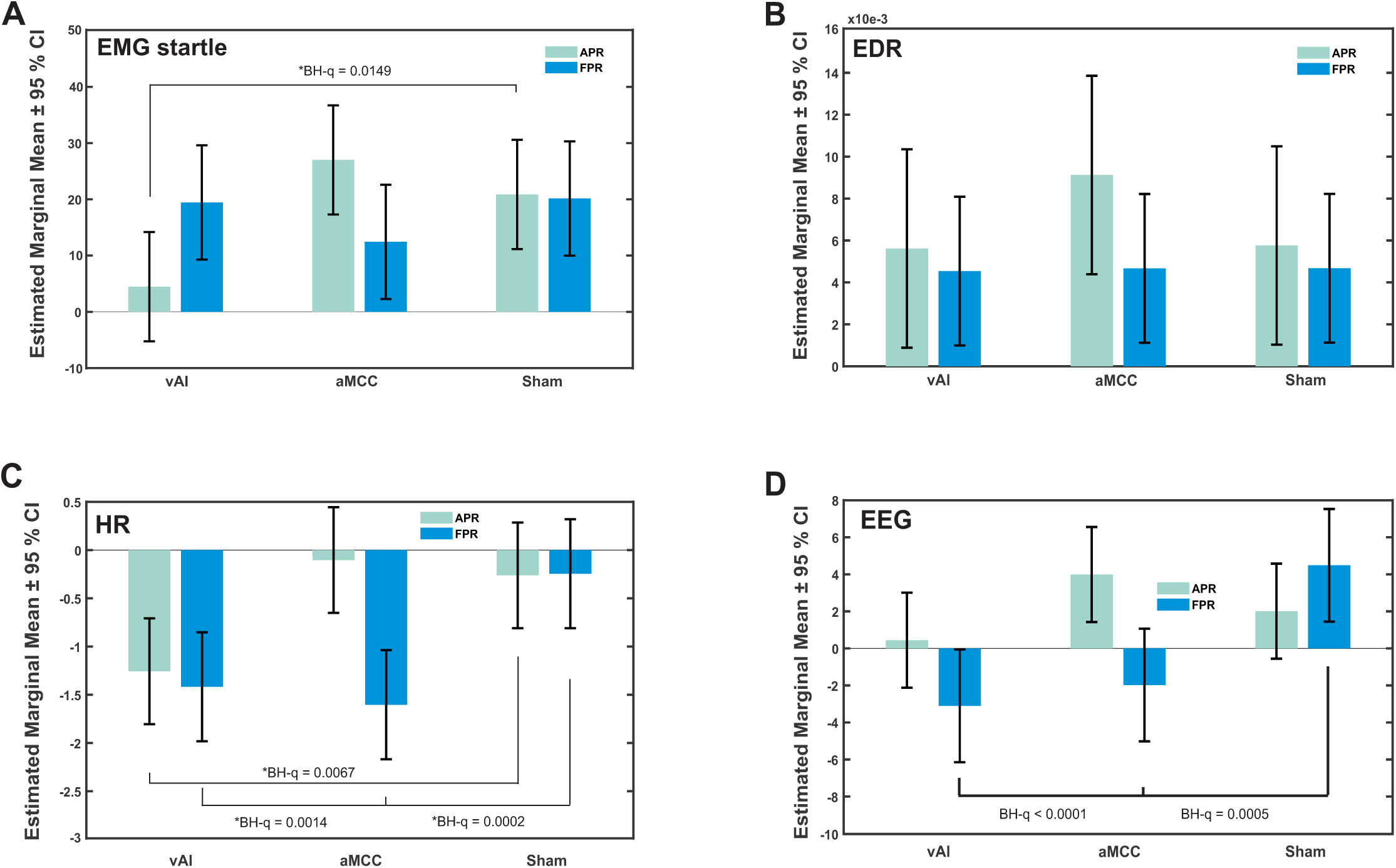
Physiological effects of LIFU on anxiety-potentiated (APR) and fear-potentiated (FPR) responding. **A.** *EMG Startle*. Bars show estimated marginal means (EMMs) ± 95% confidence intervals from the linear mixed-effects model (VAS ∼ LIFU × TASK + (1+TASK|Subject)). The model estimates indicate how LIFU (vAI, ACC) compares to Sham for each task condition, with error bars reflecting population-level uncertainty. *BH-q denotes Benjamini-Hochberg adjusted p-values. **B.** *Electrodermal Response*; **C.** *Mean Heart-Rate*; **D.** *EEG evoked potential*.

#### Electrodermal Response (EDR)

The mean ± SEM EMG area under the curve (AUC) of the EDR response to the startle probe for APR for each of vAI, aMCC and Sham conditions was: 0.0056 ± 0.0021, 0.0091 ± 0.0020 and 0.0058 ± 0.0030. For FPR, the mean ± SEM for each of vAI, aMCC and Sham conditions was: 0.0045 ± 0.0015; 0.0047 ± 0.0019 and 0.0047 ± 0.0021. All values are in arbitrary units (AU). Raw mean ± SEM are in **Supplementary Table 4 & Supplementary** Figure 5. The model showed a significant intercept (F(1,87.4) = 5.75, p = 0.019), indicating that average EDR responses were greater than zero, but there were no significant main effects of LIFU (F(2,200) = 1.33, p = 0.267), TASK (F(1,145.4) = 0.16, p = 0.687), or their interaction (F(2,200) = 0.64, p = 0.528). Planned contrasts confirmed the absence of LIFU effects. During APR, neither AI (est = –0.14, SE = 2.44, t(234) = –0.06, p = 0.953, q = 0.953) nor aMCC stimulation (est = 3.36, SE = 2.44, t(234) = 1.38, p = 0.169, q = 0.169) differed significantly from Sham. Similarly, during FPR, both vAI (est = – 0.13, p = 0.956) and aMCC (est = –0.01, p = 0.998) did not differ from Sham. Estimated marginal means illustrated this null pattern: Sham responses were comparable across APR (M = 5.76, 95% CI [1.03, 10.49]) and FPR (M = 4.67, 95% CI [1.13, 8.22]). AI stimulation produced nearly identical responses (APR M = 5.62; FPR M = 4.54), while aMCC stimulation numerically increased APR responses (M = 9.12, 95% CI [4.39, 13.86]) but not FPR (M = 4.67, 95% CI [1.12, 8.22]), though these differences were not statistically reliable. Together, these results indicate that EDR responses were not significantly modulated by LIFU or task condition (**Figure 5B**).

#### Heart Rate (HR)

The mean ± SEM HR for APR for each of AI, aMCC and Sham conditions was: -1.26 ± 0.28, -0.10 ± 0.33 and -0.26 ± 0.17. For FPR, the mean ± SEM for each of AI, aMCC and Sham conditions was: - 1.41 ± 0.33; -1.60 ± 0.25 and -0.24 ± 0.32. All values are in beats-per-minute (BPM). Raw mean ± SEM are in **Supplementary Table 5 & Supplementary** Figure 6. The model showed a significant main effect of LIFU (F(2,200) = 5.91, p = 0.0032), no main effect of TASK (F(1,199.4) = 0.00, p = 0.963), and a significant LIFU × TASK interaction (F(2,200) = 5.20, p = 0.0063). Planned contrasts indicated lower HR vs Sham under several conditions: during APR, vAI reduced HR (est = –0.995, SE = 0.364, t(234) = –2.74, p = 0.0067, BH-q = 0.0134), while aMCC did not differ from Sham (est = 0.159, p = 0.663). During FPR, both vAI (est = –1.172, SE = 0.364, t(234) = –3.22, p = 0.0014, BH-q = 0.0014) and aMCC (est = –1.358, SE = 0.364, t(234) = –3.73, p = 0.0002, BH-q = 0.0005) produced significantly lower HR than Sham. Estimated marginal means reflected this pattern: Sham HR was near zero in both APR (M = –0.262, 95% CI [–0.810, 0.287]) and FPR (M = –0.245, 95% CI [–0.810, 0.320]). vAI showed reduced HR in APR (M = –1.257, 95% CI [–1.805, –0.708]) and FPR (M = –1.417, 95% CI [–1.982, –0.852]). aMCC was similar to Sham in APR (M = –0.103, 95% CI [–0.651, 0.445]) but showed a clear reduction in FPR (M = –1.603, 95% CI [–2.168, –1.038]). Together, these results indicate that vAI LIFU reduces HR in both APR and FPR, whereas aMCC LIFU selectively reduces HR during FPR (**Figure 5C**).

#### EEG Startle Response

The mean ± SEM EEG amplitude for APR for each of vAI, aMCC and Sham conditions was: 0.44 ± 1.08, 3.99 ± 1.21 and 2.01 ± 1.15. For FPR, the mean ± SEM for each of vAI, aMCC and Sham conditions was: -3.10 ± 2.33; -1.98 ± 1.13 and 4.49 ± 1.38. All values are in microvolts (µV). Raw mean ± SEM are in **Supplementary Table 6 & Supplementary Figure 7**. The model showed there were no main effects of LIFU (F(2,200) = 1.88, p = 0.156) or TASK (F(1,137.37) = 1.44, p = 0.233), but a significant LIFU × TASK interaction (F(2,200) = 5.62, p = 0.0042). Planned contrasts showed selective reductions during FPR relative to Sham: vAI LIFU decreased EEG (est = –3.00, SE = 0.19, t(234) = –4.14, p < 0.0001, BH-q = 0.0001) and aMCC LIFU also decreased EEG (est = -2.00, SE = 0.19, t(234) = –3.52, BH-q = 0.0005). During APR, neither vAI (est = 0.20, p = 0.394) nor aMCC (est = 4.00, p = 0.281) differed from Sham. Estimated marginal means were consistent: Sham was positive in APR (M = 2.0, 95% CI [–0.56, 4.58]) and FPR (M = 4.49, 95% CI [1.45, 0.75]). vAI was near zero in APR (M = 0.44, 95% CI [–0.212, 0.301]) but negative in FPR (M = –3.10, 95% CI [–0.615, –0.062]). aMCC was positive in APR (M = 3.99, 95% CI [1.43, 0.66]) and near zero/negative in FPR (M = –1.98, 95% CI [–0.502, 0.106]). Together, these results indicate a task-dependent LIFU effect on EEG, with both vAI and aMCC reducing EEG during FPR but no APR differences versus Sham (**Figure 5D**).

### Brain-Body-Behavior Coupling

#### EMG Startle Response

In the sham-subtracted models, fluctuations in EMG startle did not significantly covary with anxiety ratings. Across conditions, the slope relating within-person EMG variation to subjective reports was not different from zero (β = 0.23, SE = 0.33, F(1,52.19) = 0.49, p = 0.48), and there were no interactions with task or stimulation site (all p > 0.34). Task-specific slopes were likewise nonsignificant: during APR, slopes were small and positive under both vAI (β = 0.23, p = 0.48) and aMCC (β = 0.29, p = 0.38) (**Figure 6A**). During FPR, slopes were negligible for vAI (β = 0.16, p = 0.49) and nonsignificant but trending negative under aMCC (β = –0.39, p = 0.11) (**Figure 6A**). Estimated marginal means (evaluated at each participant’s average EMG) indicated consistent reductions in EMG relative to Sham across conditions. vAI LIFU decreased APR startle (M = –1.02, 95% CI [–1.62, –0.42]) and FPR startle (M = –0.84, 95% CI [–1.33, –0.35]). aMCC LIFU produced a weaker, nonsignificant reduction during APR (M = –0.40, 95% CI [–1.00, 0.21]) and a reduction during FPR (M = –1.09, 95% CI [–1.59, –0.60]). Direct comparisons between vAI and aMCC revealed that vAI suppressed APR startle more strongly than aMCC (β = 0.63, SE = 0.19, p = 0.0015), whereas differences during FPR were smaller and did not reach significance (p = 0.07). Together, these results show that vAI and aMCC both reduced EMG responses relative to Sham, but only vAI produced a robust suppression during APR. Importantly, there was no evidence that variability in EMG responses to the startle probe predicted anxiety ratings, suggesting that LIFU influenced the mean magnitude of the startle reflex rather than its coupling with subjective anxiety (see **Supplemental Data** for full model results).

**Figure 6.**
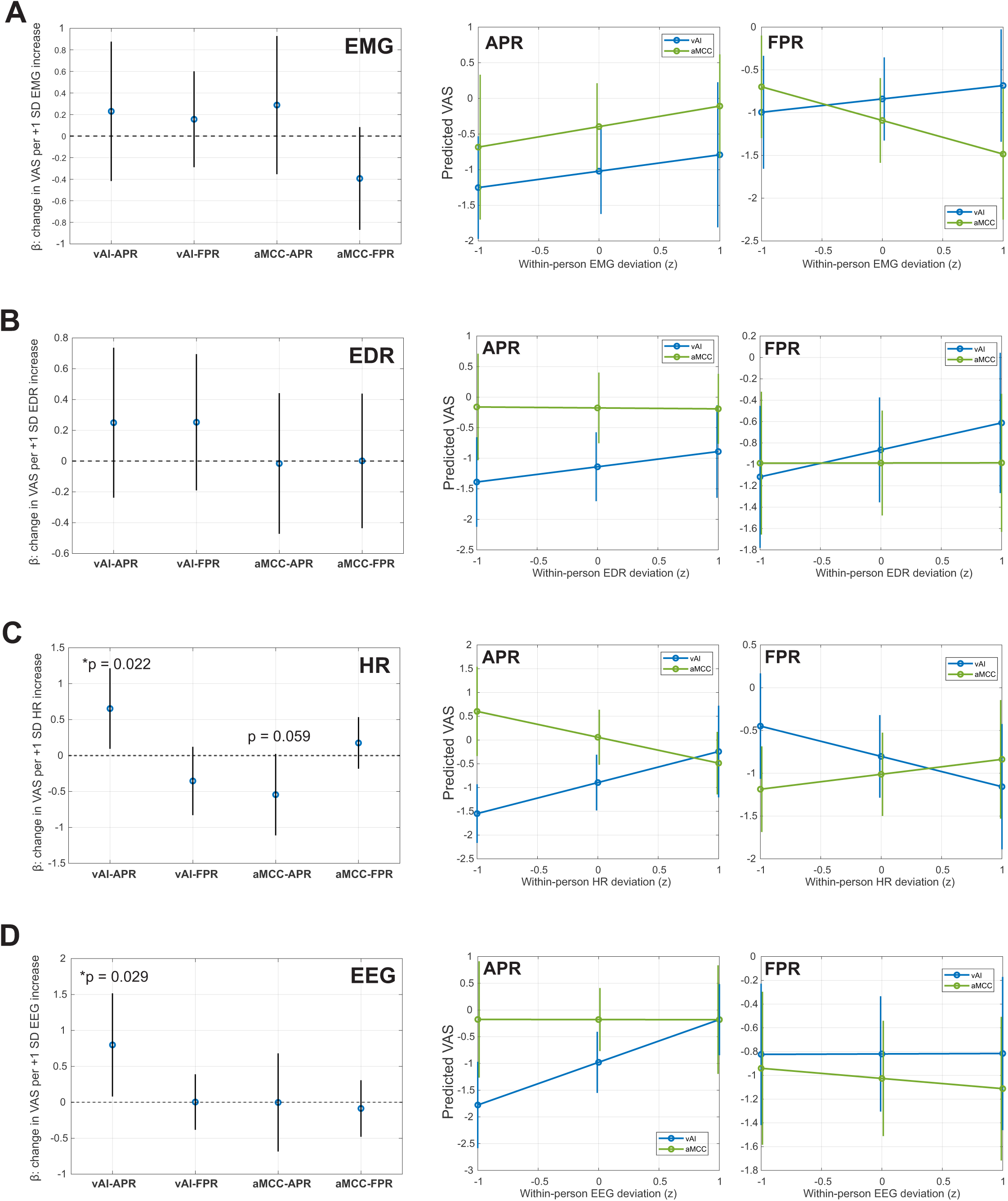
Brain-Body-Behavior Coupling. **A.** *Electromyographic (EMG) Startle Response*. (Left) LIFU (vAI, aMCC) x TASK (APR, FPR) predicted VAS change per +1 SD unit of EMG deviation. Each point is the estimated regression coefficient (β). The error bar is the 95% CI. None of the slopes were statistically significant (p > 0.11) indicated by CI bars overlapping with 0 (dashed horizontal line). (Right) Predicted VAS as a function of within-person EMG deviations by LIFU neuromodulation site for APR and FPR. Predicted APR and FPR VAS ratings are plotted against within-person deviations in EMG amplitude (z-scored) from -1 SD to +1 SD. **B.** *Electrodermal Response (EDR).* (Left) LIFU x TASK predicted VAS change per +1 SD unit of EMG deviation. Each point is the estimated regression coefficient (β). The error bar is the 95% CI. None of the slopes were statistically significant (p > 0.26) indicated by CI bars overlapping with 0 (dashed horizontal line). (Right) Predicted VAS as a function of within-person EMG deviations by LIFU neuromodulation site for APR and FPR. Predicted APR and FPR VAS ratings are plotted against within-person deviations in EMG amplitude (z-scored) from -1 SD to +1 SD. **C.** *Heart Rate (HR).* (Left) LIFU x TASK predicted VAS change per +1 SD unit of HR deviation. Each point is the estimated regression coefficient (β). The error bar is the 95% CI. The slope for LIFU to AI for the APR TASK was statistically significant (β = 0.65, t(152) = 2.131, p = 0.022). The slope for LIFU to ACC for the APR TASK was close (β = –0.55, t(152) = -1.90, p = 0.059). (Right) Predicted VAS as a function of within-person EMG deviations by LIFU neuromodulation site for APR and FPR. Predicted APR and FPR VAS ratings are plotted against within-person deviations in mean HR (z-scored) from -1 SD to +1 SD. **D.** *Electroencephalography (EEG)* (Left) LIFU x TASK predicted VAS change per +1 SD unit of HR deviation. Each point is the estimated regression coefficient (β). The error bar is the 95% CI. The slope for LIFU to AI for the APR TASK was statistically significant (β = 0.65, t(152) = 2.131, p = 0.022). The slope for LIFU to ACC for the APR TASK was close (β = –0.55, t(152) = -1.90, p = 0.059). (Right) Predicted VAS as a function of within-person EMG deviations by LIFU neuromodulation site for APR and FPR. Predicted APR and FPR VAS ratings are plotted against within-person deviations in mean HR (z-scored) from -1 SD to +1 SD.

#### Electrodermal Responses (EDR)

The sham-subtracted models revealed no evidence that within-person fluctuations in EDR covaried with anxiety ratings. Across participants, the slope relating EDR variation to symptom reports was not significant (β = 0.25, SE = 0.25, F(1,38.9) = 1.03, p = 0.31), and there were no interactions with task or LIFU condition (all p > 0.31). Task-specific slopes were similarly nonsignificant: during APR, the slope was small and positive under vAI (β = 0.25, p = 0.31) and near zero under aMCC (β = –0.02, p = 0.95); during FPR, slopes were also negligible under both AI (β = 0.25, p = 0.26) and aMCC (β = 0.00, p = 0.99) (see **Figure 6B**). The estimated marginal means (evaluated at each participant’s average EDR) showed that both vAI and aMCC LIFU produced reductions in sham-subtracted EDR during FPR (vAI: M = –0.86, 95% CI [–1.35, –0.37]; aMCC: M = –0.99, 95% CI [–1.48, –0.50]), whereas only vAI affected EDR during APR (AI: M = –1.14, 95% CI [–1.70, –0.58]; aMCC: M = –0.18, 95% CI [–0.75, 0.40]). A direct comparison between vAI and aMCC showed a reliable difference at the APR level (β = 0.96, SE = 0.17, p < 0.001), but no differences during FPR or in trial-to-trial coupling slopes. Together, these results indicate that EDR did not exhibit reliable brain–body–behavior coupling, and LIFU did not significantly alter the link between EDR fluctuations and anxiety symptoms. However, vAI produced consistently lower sham-subtracted EDR levels relative to aMCC during APR, suggesting some condition-specific divergence at the mean level rather than in coupling (see **Supplemental Data** for full model results).

#### Heart Rate (HR)

In the sham-subtracted models, heart rate showed the most consistent evidence of brain– body–behavior coupling. Across conditions, within-person increases in HR were associated with higher anxiety symptoms (β = 0.65, SE = 0.28, F(1,55.69) = 5.329, p = 0.023), indicating that participants who exhibited greater elevations in HR during the task relative to their own baseline also reported more anxiety. This effect was primarily evident during APR for LIFU to vAI (β = 0.65, t(152) = 2.131, p = 0.022), with a trend in the opposite direction under aMCC (β = –0.55, t(152) = -1.90, p = 0.059) (**Figure 6C**). No reliable associations were observed during FPR (AI β = –0.35, p = 0.14; aMCC β = 0.18, p = 0.34). Notably, the slopes differed significantly across tasks: under vAI LIFU, coupling was stronger in APR than FPR (β = –1.01, t(152) = -2.66, p = 0.0086), whereas under aMCC the reverse pattern emerged, with stronger coupling in FPR than APR (β = 1.73, t(152) = 2.71, p = 0.0075). Direct comparisons between LIFU sites confirmed that vAI produced more positive coupling slopes than aMCC in APR (β = –1.20, t(152) = -2.44, p = 0.0157). The estimated marginal means confirmed that stimulation also modulated overall HR responses. LIFU to vAI reduced HR relative to Sham during both APR (M = –0.90, 95% CI [–1.48, –0.31]) and FPR (M = –0.80, 95% CI [–1.29, –0.32]).

aMCC produced a marked HR reduction during FPR (M = –1.01, 95% CI [–1.50, –0.53]) but showed no reliable effect during APR (M = 0.06, 95% CI [–0.52, 0.64. Together, these findings indicate that vAI LIFU not only reduced HR on average but also strengthened alignment between HR and subjective anxiety during APR, whereas aMCC selectively reduced HR during FPR with marginal coupling effects (see **Supplemental Data** for full model results).

#### EEG Startle Response

In the sham-subtracted models, EEG activity showed partial evidence of covariation with anxiety ratings. Across conditions, within-person increases in EEG were associated with higher anxiety symptoms (β = 0.80, SE = 0.36, p = 0.03), suggesting that stronger neural responses were linked to heightened anxiety. This effect was task- and site-specific: during APR, coupling between EEG and anxiety ratings was significant under vAI stimulation (β = 0.80, t(152) = 2.20, p = 0.029) but absent under aMCC (β ≈ 0, p = 0.99) **(Figure 6D)**. In the FPR condition, slopes were negligible for both vAI (β ≈ 0, p = 0.98) and aMCC (β = –0.09, p = 0.67). The difference in slopes between APR and FPR under vAI trended negative (β = –0.80, p = 0.054), suggesting weaker coupling during FPR, though this did not reach statistical significance. The estimated marginal means indicated clear stimulation effects on overall EEG amplitudes. vAI reduced EEG responses relative to Sham across both APR (M = –0.98, 95% CI [–1.55, –0.41]) and FPR (M = –0.82, 95% CI [–1.30, –0.33]). aMCC also reduced FPR responses (M = –1.03, 95% CI [–1.51, –0.54]) but showed no reliable change during APR (M = –0.18, 95% CI [–0.77, 0.41]). Direct comparisons revealed that vAI suppressed APR responses more strongly than aMCC (β = 0.80, SE = 0.16, p < 0.001), while differences in FPR were small and nonsignificant (p = 0.15). Together, these results suggest that LIFU to the vAI not only reduced EEG brain responses but also enhanced the alignment between neural activity and anxiety ratings during APR, whereas aMCC primarily reduced FPR amplitudes without altering EEG–anxiety coupling (see **Supplemental Data** for full model results).

A summary of LIFU mean effects and coupling is provided in **Table 1**.

**Table 1.** Summary of the mean effects and brain-body coupling from LIFU to AI or aMCC

|  | Mean Effects |  |  |  | Coupling |  |  |  |
| --- | --- | --- | --- | --- | --- | --- | --- | --- |
|  | AI |  | aMCC |  | AI |  | aMCC |  |
|  | APR | FPR | APR | FPR | APR | FPR | APR | FPR |
| Behavior | ↓ | ↓ | -- | ↓ |  |  |  |  |
| EMG | ↓ | -- | -- | -- | -- | -- | -- | -- |
| EDR | -- | -- | -- | -- | -- | -- | -- | -- |
| HR | ↓ | ↓ | -- | ↓ | ↑ | -- | -- | -- |
| EEG | -- | ↓ | -- | ↓ | ↑ | -- | -- | -- |

Finally, we were interested to understand if baseline anxiety state or physiology as measured by mean heart-rate or HRV mediated these task-based effects.

### Baseline Effects

#### Effect of baseline anxiety severity (STAI-S & STAI-T)

We asked if baseline STAI-T or within-subject mean or session specific STAI-S predicted APR or FPR VAS ratings or predicted the effects of LIFU. The linear mixed-effects models demonstrated neither trait or state anxiety explained variability in APR/FPR anxiety ratings or altered the effect of vAI/aMCC LIFU. Baseline anxiety did not predict APR or FPR and did not moderate LIFU effects. In mixed-effects models with random subject intercepts, LIFU alone did not predict either outcome (APR: F(2,78)=0.77, p=.468; FPR: F(2,78)=0.57, p=.570). Adding trait anxiety (STAI-T) or state anxiety decomposed into between-person and within-person components did not improve model fit either for APR (LR χ²(1)=2.66, p=.103; LR χ²(2)=2.85, p=.241) or FPR (LR χ²(1)=3.18, p=.074; LR χ²(2)=3.38, p=.184). All STAI × LIFU interactions were non-significant for both outcomes (all p ≥.34). Results were unchanged with random slopes for STAI-S-within and when residualizing STAI-S-between on STAI-T.

To test whether anxiety level moderated LIFU effects on behavior, we formed Low/High groups via median splits on STAI-T at the subject level, on STAI-S computed as each subject’s between-person mean across sessions, and on session-specific STAI-S within each LIFU session (vAI, aMCC, Sham), assigning ties to High. For APR and FPR separately, we fit linear mixed-effects models with fixed effects for LIFU, Anxiety Group, and their interaction and a random intercept for Subject (Satterthwaite degrees of freedom), and we ran planned difference-of-differences (DoD) contrasts to test whether LIFU reduced outcomes more in the High than the Low group (aMCC–Sham and AI–Sham). The STAI-T median was 39.5 and the subject mean STAI-S median was 29.2. The STAI-S session medians and counts (Low/High) were: vAI = 27.5 (20/20); aMCC = 28 (19/21); Sham = 29.5 (20/20). See **Figure 3A**. For STAI-T, APR showed a significant LIFU main effect (p=0.0045) but no Group effect (p=0.891) or interaction (p=0.774); contrasts were non-significant (aMCC–Sham p=0.557; vAI– Sham p=0.952). FPR likewise showed a LIFU main effect (p=0.0010) without Group (p=0.952) or interaction (p=0.861); contrasts were non-significant (p=0.604 and p=0.685). For between-person mean STAI-S, APR again showed a LIFU main effect (p=0.00080) but no Group (p=0.855) or interaction (p=0.649), with non-significant contrasts (p=0.382, p=0.479); FPR mirrored this pattern (LIFU p=0.00121; Group p=0.670; interaction p=0.890; contrasts p=0.699, p=0.657). Session-wise STAI-S yielded the same conclusion: APR (LIFU p=0.000634; Group p=0.761; interaction p=0.773; contrasts p=0.612, p=0.487) and FPR (LIFU p=0.005802; Group p=0.864; interaction p=0.929; contrasts p=0.986, p=0.750). Collectively, these analyses show robust LIFU main effects on APR and FPR but no evidence that trait or state anxiety groups differ in baseline APR/FPR or that anxiety moderated the behavioral impact of LIFU.

#### Baseline Physiological Predictors and LIFU moderation of APR and FPR

Because autonomic tone relates to anxiety and could plausibly shape responsiveness to neuromodulation, we first quantified associations between baseline HR/HRV and anxiety (STAI-T and session mean STAI-S) and tested within-person covariation across sessions.

#### Baseline HRV (RMSSD)

RMSSD is the root-mean square of successive differences between adjacent NN intervals that may preferentially reflect high-frequency vagal (parasympathetic) activity that has previously been associated with anxiety and may index an endophenotype of pathological anxiety (79,80). As such, we examined how and if baseline RMSSD associated with anxiety severity using the STAI-T and averaged session STAI-S. Across participants RMSSD was not related to mean STAI-S (r = -0.248, p = 0.124) and trended to be inversely related to STAI-T (r = -0.303, p = 0.057) (see **Figure 7A**). We next asked whether baseline RMSSD predicts APR or FPR VAS ratings and also LIFU responsiveness. Using the moderation model (APR ∼ LIFU + HRV-between × LIFU + HRV-within × LIFU + (1|subject)), neither between-person nor within-person RMSSD predicted APR (HRV-between: F(1,119.7)=0.58, p=0.449; HRV-within: F(1,118.0)=0.35, p=0.553), and neither interaction with LIFU was significant (HRV-between × LIFU: F(2,102.5)=0.28, p=0.758; HRV-within × LIFU: F(2,115.7)=0.043, p=0.958). The main effect of LIFU was not significant. For FPR, RMSSD effects and RMSSD × LIFU interactions were also non-significant (all p≥0.54). However, state anxiety covaried within-person with RMSSD across sessions (STAI-S ∼ RMSSD-within: F(1,120)=4.24, p=0.0416) (**Figure 7A**), whereas a binary above-own-mean RMSSD did not reach significance (F(1,120)=2.98, p=0.087). Thus, the correlations demonstrate that people with lower average RMSSD have higher average STAI-S and the within-person effect shows that on days when a given person’s RMSSD is above their own average, their STAI-S is correspondingly lower (or vice versa) (see **Figure 7A**).

**Figure 7.**
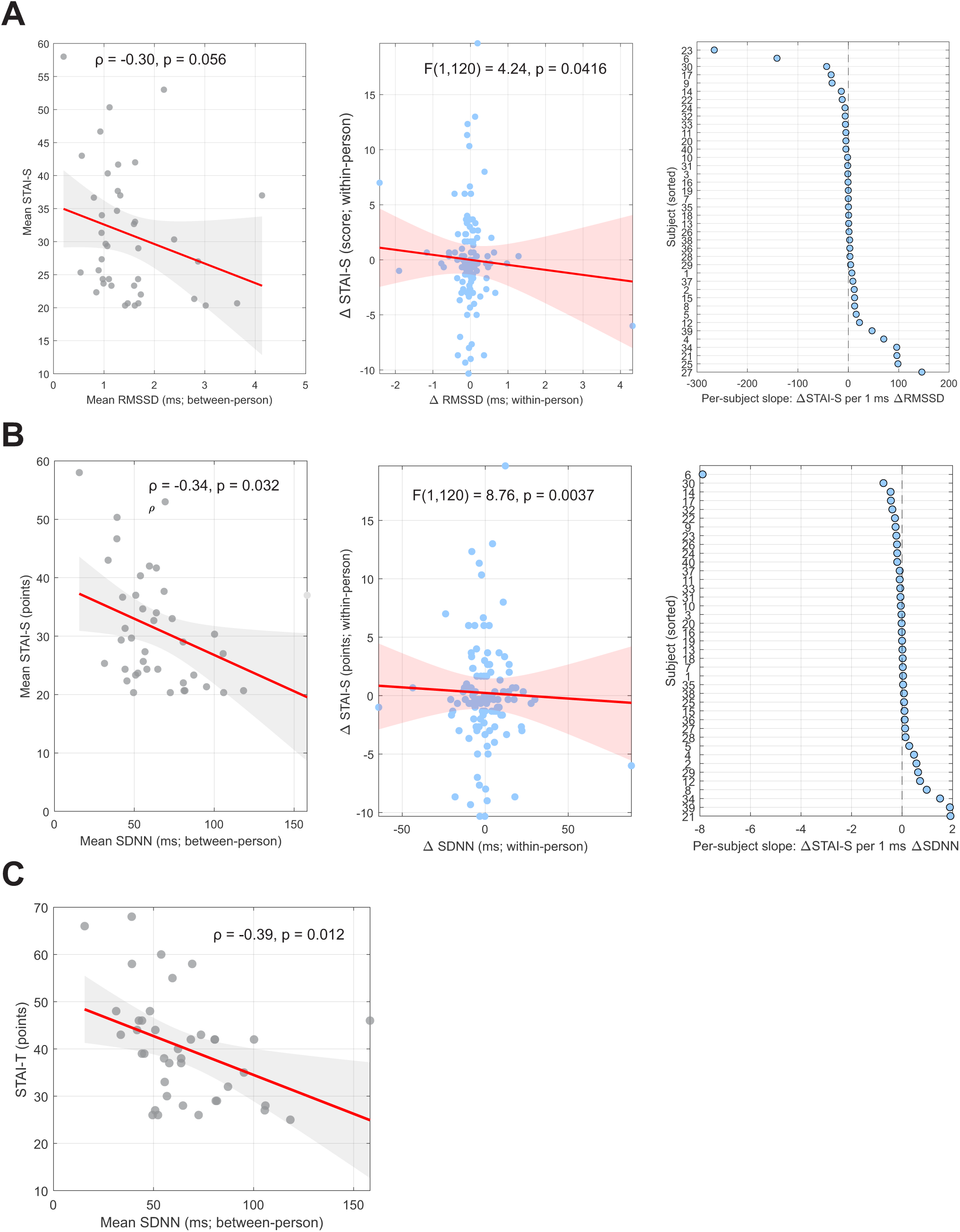
HRV & State Anxiety Coupling. RMSSD. A. (Left). *Between-person correlation*. Average RMSSD across sessions plotted against average STAI-S. Between-person association using each participant’s mean RMSSD versus mean STAI-S. Each gray circle is one participant. The solid red line is the OLS fit with gray 95% CI band. In contrast to the within-person result, the between-person relationship is weaker (Spearman ρ≈−0.30, p≈0.056), highlighting that the primary effect detected by the LMM is within individuals over time rather than differences between individuals. **(Middle).** *Within-person coupling*. Session-level deviations in RMSSD (ΔRMSSD) relative to each participant’s mean predicted concurrent deviations in STAI-S (ΔSTAI-S). Each light-blue circle is one session from one participant (N = 120 session-level observations; 40 participants × 3 sessions: AI, aMCC, Sham). The x-axis shows ΔRMSSD (ms), computed as each session’s RMSSD minus that participant’s own mean. The y-axis shows ΔSTAI-S centered within person. The solid red line is the marginal population slope from the linear mixed model (LMM: ΔSTAI-S ∼ 1 + ΔRMSSD + Session + (1|Subject)), averaged across sessions (AI/aMCC/Sham). The red band is the 95% CI. The within-person effect is significant (F(1,120)=4.24, p=0.0416; partial η²≈0.034), indicating that days when a participant’s RMSSD deviates from their own average are associated with concurrent deviations in their STAI-S in the direction of the slope. **(Right).** Subject-wise ordinary least-squares (OLS) slopes of ΔSTAI-S vs ΔRMSSD, showing heterogeneity in within-person associations around the population effect. Subject-wise within-person slopes are shown as light-blue circles, one point per participant, sorted from most negative to most positive. Each slope is the ordinary-least-squares line fitted to that person’s three centered sessions (ΔRMSSD vs ΔSTAI-S). The vertical black dashed line at 0 marks no association. The spread of points on both sides of zero illustrates heterogeneity in individual within-person coupling: Some participants show stronger negative coupling, others weak or positive coupling. **B. (Left).** *Between-person correlation*. Average SDNN across sessions plotted against average STAI-S. Each gray circle is one participant. The solid red line is the ordinary-least-squares fit with a gray 95 % CI band. The between-person correlation is inverse and statistically significant by rank-based analysis (Spearman ρ = −0.34, p = 0.032), consistent with higher baseline SDNN associating with lower mean state anxiety. **(Middle).** *Within-person coupling*. Session-level deviations in SDNN (ΔSDNN) relative to each participant’s mean predict concurrent deviations in STAI-S (ΔSTAI-S), with a significant negative slope.Panel A (left): Each light-blue filled circle is one session from one participant (N = 120 session-level observations; 40 participants × 3 sessions: AI, aMCC, Sham). The x-axis shows ΔSDNN (ms), computed as the session’s SDNN minus that participant’s own mean. The y-axis shows ΔSTAI-S centered within person. The solid red line is the marginal population slope from the linear mixed model (ΔSTAI-S ∼ 1 + ΔSDNN + Session + (1|Subject)), averaged across sessions. The red band is the 95% CI. The within-person effect is significant (F(1,120)=8.76, p=0.0037), indicating that days when a participant’s SDNN is above their own average are associated with corresponding deviations in their state anxiety in the direction indicated by the red slope. **(Right).** *Individual Slopes.* Subject-wise OLS slopes of ΔSTAI-S vs ΔSDNN, showing within-person associations around the population effect. Subject-wise within-person slopes are shown as light-blue circles, one per participant, sorted from most negative to most positive. The vertical black dashed line at 0 marks no association. The spread of points around zero illustrates heterogeneity in individual within-person coupling. **C.** Between-person association of SDNN with trait anxiety (STAI-T). Each gray circle is one participant’s mean SDNN plotted against their STAI-T score. The solid red line is the ordinary-least-squares fit, and the gray band is the 95% CI. The association is inverse and statistically significant (e.g., Spearman ρ = −0.39, p = 0.012), indicating that participants with higher baseline SDNN tend to report lower trait anxiety.

**Figure 8.**
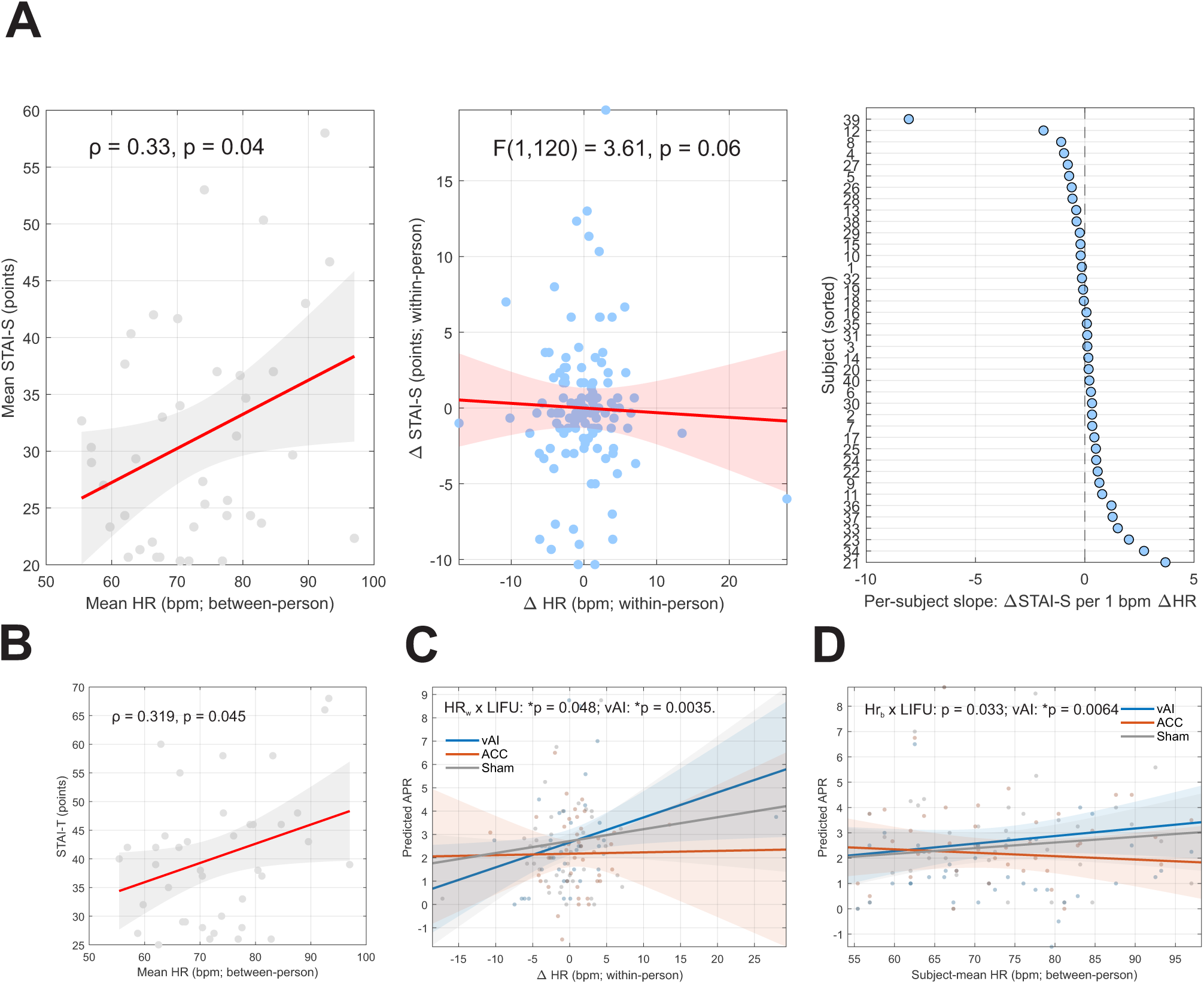
HR & State Anxiety Coupling. A. (Left). *Within-person coupling*. Session-level deviations in mean HR (ΔHR) relative to each participant’s mean predict concurrent deviations in STAI-S (ΔSTAI-S), with a significant negative slope. Each light-blue filled circle is one session from one participant (N = 120 session-level observations; 40 participants × 3 sessions: AI, aMCC, Sham). The x-axis is ΔHR (bpm), computed as the session’s heart rate minus that participant’s own mean. The y-axis is ΔSTAI-S centered within person. The solid red line is the marginal population slope from the linear mixed model (ΔSTAI-S ∼ 1 + ΔHR + Session + (1|Subject)), averaged over session levels. The red band is the 95% CI. Within-person covariation is trend-level (F(1,120)=3.61, p=.060): Days with above-mean HR tended to coincide with higher STAI-S. **(Middle).** *Individual Slopes*. Subject-wise OLS slopes of ΔSTAI-S vs ΔRMSSD around the population effect. Subject-wise within-person slopes (ΔSTAI-S per 1 bpm ΔHR) appear as light-blue circles, one per participant, sorted from most negative to most positive. The vertical dashed line at 0 marks no association. The spread around zero shows heterogeneity in individual within-person coupling. **(Right).** *Between-person correlation*. Average HR across sessions plotted against average STAI-S. Between-person association of mean HR (bpm) vs mean STAI-S. Each gray filled circle is one participant. The solid red line is the OLS fit with gray 95% CI band. The between-person correlation is positive and significant (Spearman ρ = 0.327, p = 0.040), indicating that participants with higher baseline heart rate report higher average state anxiety. In OLS scaling, a +5 bpm increase in resting HR predicts ≈ +1.5 STAI-S points (95% CI +0.08 to +2.93, p = .04). **B. (Left).** *Between-person association of heart rate with trait anxiety (STAI-T).* Each gray filled circle is one participant’s mean HR (bpm) plotted against STAI-T score. The solid red line is the OLS fit; the gray band is the 95% CI. The association is positive and statistically significant (e.g., Spearman ρ = 0.319, p = 0.045). Participants with higher baseline heart rate tend to report higher trait anxiety. **C***. Moderation of APR by baseline heart rate*: *Within-person moderation*. Lines show predicted APR as a function of ΔHR (bpm; within-person deviation from each participant’s mean) with HR-between fixed at 0 (participant mean HR), plotted separately by LIFU condition: AI (blue), aMCC (orange), Sham (gray). Shaded ribbons are 95% CIs. The HR-within × LIFU interaction is significant (F(2,111.9)=3.12, p=.048). Simple-slope tests reveal a positive HR–APR slope under AI (p=.0035). **D.** *Between-person moderation*. Lines show predicted APR as a function of subject-mean HR (bpm; between-person component) with HR-within fixed at 0 (participant mean day), by LIFU (AI blue, aMCC orange, Sham gray) with 95% CI ribbons. The HR-between × LIFU interaction is significant (F(2,98.7)=3.54, p=.033), driven by a positive association under AI (p=.0064).Together, **C & D** illustrate that baseline heart rate moderates AI-specific APR changes at both within-person (day-to-day fluctuations) and between-person (trait-like mean HR) levels.

#### Baseline HRV (SDNN)

SDNN is the standard deviation of NN (normal-to-normal) intervals. SDNN is complementary to RMSSD as it looks to reflect total variability across both low and high frequency HRV bands(81,82). Repeating the above analysis with baseline SDNN we found statistically significant inverse relationship of SDNN with mean STAI-S (ρ = -0.34, p = 0.032) and with STAI-T (ρ = -0.39, p = 0.012) (**Figure 7B & C**). Thus, across participants, higher baseline SDNN was associated with lower anxiety. In ordinary least squares (OLS) models using subject-mean SDNN (z-scored), each + 10 msec increase in SDNN predicted - 1.24 points in mean STAI-S (95% CI [-2.37 -0.11], p = 0.032) and -1.65 points in STAI-T (95% CI [-2.91 -0.38], p = 0.012) consistent with the inverse SDNN-anxiety relationship (see **Figure 7B**). In the mixed-model analysis, we found no evidence that baseline SDNN moderates LIFU effects on APR or FPR ratings. In the APR model, neither SDNN component nor its interactions with LIFU were significant (HRV-between: F(1,119.9)=1.47, p=0.228; HRV-within: F(1,117.9)=0.00017, p=0.990; HRV-between × LIFU: F(2,99.7)=0.83, p=0.440; HRV-within × LIFU: F(2,113.6)=0.16, p=0.851). The LIFU main effect was also non-significant. FPR results were likewise null (all p≥0.53). Similar to RMSSD, SDNN showed a robust within-person association with state anxiety: STAI-S covaried with SDNN-within (F(1,120)=8.76, p=0.0037) (**Figure 7B**), and sessions with above-own-mean SDNN also differed in STAI-S (F(1,120)=9.14, p=0.0031).

#### Baseline Heart Rate

In addition to HRV, we also examined baseline heart-rate and its relationship to anxiety severity and also as a predictor of effects due to the known associations of anxiety with vagal control of the heart, heart-beat interoception and cardiovascular pathology (83–85). The Spearman correlation revealed a significant positive relationship with mean STAI-S (r = 0.327, p = 0.04) (**Figure 7A**) and a significant positive relationship with STAI-T (r = 0.319, p = 0.045) (**Figure 7B**). Across participants, higher baseline heart rate was associated with greater anxiety. In OLS models, + 5 bpm increase in resting HR predicted + 1.5 points in mean STAI-S (95% CI [+0.08, +2.93], p = 0.04) and + 1.68 points in STAI-T (95% CI [+0.04, + 3.31], p = 0.045).

These between-person effects align with the within-person trend (below) that days with above-mean HR coincided with higher STAI-S. Across sessions, STAI-S tended to be higher on days when HR exceeded a participant’s own mean (STAI-S ∼ HR-within: F(1,120)=3.61, p=.060; binary “above-mean HR”: F(1,120)=2.72, p=.10), indicating only weak within-subject covariation (**Figure 7A**). In a linear mixed-effects moderation model decomposing HR into between-person and within-person components (APR ∼ LIFU + HR-between × LIFU + HR-within × LIFU + (1|subject)), both HR components were significant (HR-between: F(1,119.9)=7.73, p=.0063; HR-within: F(1,113.4)=8.90, p=.0035) and both interacted with LIFU (HR-between × LIFU: F(2,98.7)=3.54, p=.033; HR-within × LIFU: F(2,111.9)=3.12, p=.048) (**Figure 7 C & D**). Simple-slope tests showed a significant HR–APR association under AI (within: p=.0035; between: p=.0064) with nonsignificant slopes for aMCC and Sham (p ≥ .52). The LIFU main effect was not significant. For FPR, neither HR terms nor HR × LIFU interactions were significant (all p ≥ .52).

## Discussion

Anxious states are associated with heightened brain–body reactivity to both uncertain and predictable threats, yet causal evidence for the deeper cortical hubs orchestrating these states has been limited. Guided by models of interoception and salience-network function in mental health(3,5,86), we targeted the ventral anterior insula (vAI) and anterior mid-cingulate cortex (aMCC) with low-intensity focused ultrasound (LIFU) during the NPU threat task, selecting a non-clinical sample for this initial evaluation. We hypothesized that vAI would preferentially support anticipatory anxiety under uncertainty (APR), whereas aMCC would contribute more directly to defensive mobilization in response to predictable threat cues (FPR) given that the anterior insula looks to preferably encode subjective anxious arousal and interoceptive anticipation under uncertainty and the aMCC is engaged in conflict monitoring, allocation of control and mobilization of defensive responses which may be more linked to imminent threat or fear potentiated responses (11,12). Our findings support a dissociation. LIFU to the vAI reduced both APR and FPR symptom responses, whereas LIFU to aMCC selectively reduced FPR only. Physiological measures largely paralleled behavior: LIFU to the vAI decreased APR-linked EMG startle and heart rate, whereas aMCC effects were confined to FPR, with reductions in EEG amplitude and HR. In addition, LIFU to the vAI increased the coupling of both HR and EEG with APR symptom report such that transient changes in HR and EEG response predicted concomitant changes in APR symptoms. This was not found for FPR nor did LIFU to the aMCC have any effect on this brain-body-behavior coupling.

### Behavior

APR and FPR are derived indices of modulation that capture distinct facets of threat processing(22,87). In the context of the NPU task, APR reflects heightened anticipatory anxiety when threat is uncertain (U no Cue – N no Cue), a state that has been linked to hypervigilance and intolerance of uncertainty(12,88). Given its role in interoceptive prediction processing (1), the triggering of emotional responses to stimuli (89), and involvement in autonomic control (7,114), the vAI provides a plausible neural substrate for exaggerated APR such that the environmental uncertainty alters the organism’s ability to create stable predictions and, when compared to incoming sensory signals, generates large prediction errors and resultant autonomic responses to prepare the body for action. Accordingly, our data suggest it is conceivable that LIFU to the vAI may reduce APR by inhibiting uncertainty signals and minimizing prediction errors, thus reducing physiological and subjective expressions of anticipatory anxiety. Indeed, our previous results suggest that the effect of LIFU in the human brain is consistent with inhibition (50,66,67,90,91).

Interestingly, LIFU to the vAI also reduced FPR VAS responses. While we originally hypothesized a preferential response to APR, (and directly comparing vAI and aMCC effects on APR supported this hypothesis) the effects of vAI on FPR are not surprising given the strong connectivity of the vAI with the amygdala and aMCC with both regions’ well-established roles in fear processing(15,92,93). The ventral anterior insula (vAI) is tightly coupled with the amygdala(94), particularly the central and basolateral nuclei, and these connections are bidirectional. Animal tracing and optogenetics show that the anterior agranular insula provides a major source of projections to the amygdala, and lesions or inhibition in this region reduce both anxiety-like behaviors and conditioned fear responses (95). In parallel, direct anatomical work demonstrates that the insula communicates with central amygdala output streams that control defensive physiology and behavior, including hypothalamic and brainstem targets (96). From the perspective of fear learning, large-scale human meta-analyses consistently identify the insula and amygdala as co-activated during cue-conditioned fear (97). Circuit reviews also highlight that the extended amygdala (central amygdala + bed nucleus of the stria terminalis) integrates both phasic fear and sustained anxiety, and that the insula provides key interoceptive input into this network (98,99). LeDoux’s “two-systems” framework underscores this point: while the amygdala coordinates defensive behavioral and physiological responses to immediate threats, cortical regions such as the insula shape the subjective experience of fear and anticipation (100). Recent clinical and translational studies also show that anterior insula activation tracks individual differences in fear of pain and anticipatory threat, sometimes more robustly than amygdala activity itself (101). This suggests that the vAI is not only central to anticipatory anxiety (APR) but also engaged whenever bodily threat cues are processed in concert with amygdala-driven fear.

LIFU to the aMCC exclusively affected FPR symptoms. FPR, defined as the difference between cued and un-cued predictable threat conditions, reflects cue-potentiated fear reactivity (22). Higher FPR suggests heightened reactivity to known threat cues, whereas lower FPR may reflect improved regulatory control (18,22). The reduction of FPR symptom responses was not exclusive to the aMCC but inhibition of the aMCC did not affect APR symptoms suggesting a dissociation of the role of vAI and aMCC in APR and FPR mechanisms. This was consistent with our initial hypothesis based on the aMCC role in preparation for defensive responses in the face of predictable threats. The anterior mid-cingulate cortex (aMCC; often labeled dorsal ACC(47,102)) is consistently implicated in mobilizing defensive responses to explicit, predictable threat cues. Meta-analytic and empirical data demonstrate that the aMCC is a hub where information about reinforcers and aversive stimuli is integrated with motor and autonomic output systems, supporting its role in preparing and executing defensive actions (98,103,104). Indeed, recent viral tracing work shows the aMCC has descending connections to the adrenal medulla, providing an anatomical basis for modulating sympathetic activity(105). Fear-conditioning studies and neuroimaging work highlight aMCC engagement when conditioned cues predict imminent threat, distinguishing its function from more ventral cingulate regions involved in regulation (16,93,106). This fits with threat imminence models in which the aMCC becomes increasingly engaged as danger is close and requires rapid, organized responding (55). The selectivity of the aMCC for predictable threat is also supported by recent translational work. Radoman and colleagues found that predictable threat robustly activated the aMCC along with thalamus and brainstem, whereas unpredictable threat more strongly recruited the anterior insula (107). A companion study further suggested that temporally unpredictable threat engages both anterior insula and aMCC, but with stronger differentiation of aMCC during threat relative to uncertainty, underscoring its role in phasic, cue-driven responding (108). From a clinical perspective, exaggerated aMCC activity has been observed across fear-based anxiety disorders during cue-provoked fear, but is less consistently engaged in sustained anxiety or intolerance of uncertainty (15,106,108). This dovetails with the present findings that LIFU to the aMCC reduced fear-potentiated responses (FPR), which reflects cue-driven reactivity to predictable threat, while leaving anxiety-potentiated responses (APR) unaffected.

#### Physiological Markers

##### Heart-rate

The univariate analysis showed vAI LIFU-associated reductions in heart-rate for APR and FPR. The link between heart rate and mental health and anxiety is well established (5,30,34) and the link between anxiety and cardiovascular disease is also established(83) with evidence that exercise can lower HR and improve responsiveness during threat tasks(109). De Pascalis et al. found that higher trait anxiety is associated with poorer perception of cardiac activity, suggesting anxiety disrupts cardiac interoceptive accuracy(110). Garfinkel et al. extended this, showing cardiac cycle timing gates fear perception such that fearful faces presented at systole were rated as more intense and evoked stronger amygdala responses, directly linking cardiac afferents to fear appraisal (111). The effect on HR during a threat task is supported by several lines of evidence that link the insula, anterior cingulate cortex and amygdala as a core cortical autonomic network regulating cardiac function (38,112–114). The insula is central for limbic–autonomic integration(115), consistent with the finding that vAI LIFU reduced HR under APR. The aMCC, by contrast, is more tightly tied to defensive mobilization and attentional control, which could explain why HR was affected during FPR for aMCC LIFU. Meta-analytic and clinical studies consistently report elevated HR across anxiety disorders (116,117). Though not central to anxiety, Loggia et al. (118) showed HR tracks subjective pain perception, supporting HR as a sensitive marker of aversive states. Similar to the cardiac timing of fearful faces, timing of painful stimuli to systole alters the amplitude of evoked potentials and autonomic responses that correlate with insular BOLD responses (119,120). By analogy, HR reductions after vAI and aMCC LIFU could reflect down-regulation of perceived threat and/or anxiety states. Importantly, our baseline HR analyses reinforce this interpretation. Within individuals, higher baseline HR covaried with higher state anxiety (STAI-S), and mixed-effects modeling showed that HR moderated the effect of LIFU on APR but not FPR. In our brain-body-behavior coupling analyses, HR was positively coupled with APR under vAI LIFU but not under aMCC, suggesting that interindividual and intraindividual differences in cardiac state shaped the degree to which neuromodulation reduced anticipatory anxiety. This pattern is consistent with models linking interoceptive cardiovascular signals to anxious anticipation and situates the vAI as a key node where cardiac–affective integration can be down-regulated by LIFU. Our findings show that neuromodulation of vAI and aMCC can alter how baseline cardiac state interacts with anticipatory anxiety, underscoring the dynamic brain–heart axis as a mechanistic substrate of LIFU effects.

##### Startle and Evoked Potential

Our univariate analyses found LIFU to vAI reduced the EMG startle response during APR whereas EEG evoked potential response to the startle probe was only reduced during the FPR conditions for both LIFU to vAI and aMCC. Convergent NPU and startle–ERP evidence suggests that when anticipatory anxiety (APR) decreases, defensive reflexes and early cortical responses should drop in tandem— though we did not observe this. This disconnect between effects on the startle response and the EEG response may be due to the neural architecture mediating these responses. Startle is mediated by a rapid subcortical reflex arc involving the brainstem, amygdala and insula with direct projections onto motor pathways (121–123) whereas the ERP during threat probes reflect cortical processing of attention and salience recruiting aMCC and insula. This suggests that aMCC modulation might preferentially impact cortical threat appraisal (represented via the ERP responses in our study) without necessarily modulating the subcortical reflex loop underlying startle (88). In a pair of papers, Nelson et al. showed that startle and ERPs could be dissociated. They found that startle is more tightly linked to reflexive defensive responses while ERPs reflect cortical attentional allocation(24,124). Thus, it could be speculated that the aMCC is a cortical hub directly contributing to stimulus-locked ERPs but it does not strongly modulate the sub-cortical reflex arc generating the startle response. In contrast, vAI with its stronger connectivity to amygdala and brainstem influences both.

##### Electrodermal Response

We did not find any evidence for LIFU to the vAI or aMCC to affect the EDR response. The absence of LIFU effects on EDR is consistent with the roles of the vAI and aMCC as integrators of salience, interoception, and cognitive control, rather than as direct generators of sympathetic sudomotor output (125). While both regions co-activate with autonomic arousal, the actual expression of skin conductance is primarily determined by hypothalamic–brainstem pathways, with strong contextual modulation from the vmPFC in anticipatory states and the amygdala during fear. In this framework, modulating the vAI or aMCC alone would therefore be unlikely to shift the downstream gain of the sudomotor system, which helps explain why APR and FPR responses were unaffected (125). Our previous work exploring LIFU to the vAI and aMCC in the context of pain and autonomic response also did not find EDR responses (50,90).

#### Brain-body-behavior coupling

Our sham-subtracted analyses revealed a site × context double dissociation in brain–body–behavior coupling. During anticipatory anxiety (APR), ventral anterior insula (vAI) LIFU both reduced mean HR/EEG and strengthened within-person coupling between HR (and EEG) and anxiety ratings while for fear-potentiated responses (FPR), aMCC LIFU primarily lowered mean responses with weak or absent coupling effects. Direct vAI vs aMCC contrasts confirmed more positive coupling slopes under vAI in APR, and the opposite pattern under aMCC in FPR. These findings indicate that neuromodulation can reshape not only group mean response magnitude (univariate analyses) but also the alignment between physiology, neural signals, and subjective experience, in a manner that depends on both node (vAI vs aMCC) and threat context (uncertainty/APR vs cue/FPR).

This pattern fits with contemporary models of interoception and salience processing in the insula. Viscerosensory signals ascend via lamina I/spinothalamic and vagal pathways and are putatively integrated through brainstem and thalamic nuclei before projecting topographically to the dorsal posterior insula(4,126). According to the embodied predictive interoception coding (EPIC) model (1) viscerosensory signals in the PI are compared to ongoing predictions of expected bodily states generated in the anterior insula (and other regions) with the resulting prediction errors relayed back to the vAI (and other cortical areas) to initiate descending visceromotor commands. It is these descending signals that would alter peripheral physiology (i,e, heart-rate) to respond to task demands. Experimental work supports that the anterior insula (as part of a broader, distributed network) exerts beat-to-beat autonomic control of the heart via medullary and spinal circuits(38,105,114,127). This interoceptive loop, principally in the anterior insula, is hypothesized to link bodily states with affect and subjective awareness(128,129). Our directional observation of LIFU to the vAI down-regulating mean arousal yet sharpening HR/EEG to VAS coupling under uncertainty can be seen as plausible evidence supporting the EPIC model. For example, by disrupting prediction formation and visceromotor output in the vAI, LIFU attenuated neural, autonomic, and concomitant behavioral ratings to uncertain threat, but theoretically leaving ascending viscerosensory signals to the posterior insula intact.

Placing the results within the salience-network framework helps further reconcile the mean-level suppression with stronger coupling under vAI in APR responses observed in the current study. The salience network, anchored in the vAI and rostral aMCC/anterior cingulate, integrates internal (viscerosensory) and external inputs to prioritize behaviorally relevant information and coordinate large-scale network dynamics(3,9,114). The aMCC pattern of robust mean suppression with minimal coupling change in FPR aligns with its role in action selection, performance monitoring, and cue-contingent control within the fear/anxiety circuit(130).

Human and translational literatures consistently implicate insula, amygdala, hippocampus, and cingulate subregions in fear conditioning and extinction, with dACC/aMCC tracking conditioned responding and error-like signals. In predictable, cue-driven contexts (such as the FPR condition evoked in the present study), aMCC neuromodulation may therefore optimize cue-linked response control (lowering overall HR/EEG) without proportionally increasing trial-level brain–body–report alignment, consistent with weaker within-person slopes.

#### Clinical Implications

Disorders marked by aberrant interoception and salience processing e.g., anxiety phenotypes with elevated resting HR and hypervigilance show insula dysfunction and disrupted coupling of autonomic signals to conscious feeling states(131–133). The present site- and context-specific LIFU effects suggest testable therapeutic strategies: targeting vAI to down-shift arousal and restore brain–body–behavior coherence where uncertainty dominates, and targeting aMCC to tamp cue-linked hyperreactivity.

#### Baseline Effects: STAI-T and STAI-S

As an exploratory analysis we were interested to see if baseline STAI-T or STAI-S levels influenced anxiety ratings (APR, FPR) or moderated the effects of LIFU as previous research has demonstrated state anxiety levels to affect behavior in a threat task and more so for the anterior insula activity to be positively associated with state anxiety level(134). Mixed-effects models consistently indicated that neither trait nor state anxiety accounted for variability in behavioral outcomes, and including anxiety terms did not improve model fit. Moreover, no evidence emerged for STAI × LIFU interactions (as was found in (134)), suggesting that the behavioral effects of vAI and aMCC stimulation were not contingent on individual differences in anxiety. Median-split analyses reinforced these findings. Across multiple operationalizations of anxiety grouping (trait-based, person-mean state, and session-specific state), LIFU exerted significant main effects on APR and FPR, but there were no reliable group differences or interactions. Thus, participants with higher baseline anxiety responded to LIFU similarly to those with lower anxiety at least within the healthy cohort range sampled here. These results suggest that LIFU effects on anxiety-related ratings are robust and not dependent on pre-existing differences in anxiety severity. While anxiety is an important clinical phenotype, our findings indicate that baseline anxiety level neither predicts behavioral responses to LIFU nor alters its efficacy in modulating subjective anxiety during task performance. This stability may be advantageous for translational applications, as it implies that LIFU effects are generalizable across individuals with varying levels of baseline anxiety.

#### Baseline Effects: HRV

Our findings provide a nuanced view of baseline HRV (RMSSD, SDNN) in relation to anxiety severity and responsiveness to LIFU. Consistent with prior meta-analyses(79,135), we observed evidence that lower HRV is associated with higher trait and state anxiety across participants, particularly for SDNN, which showed robust inverse associations with both STAI-S and STAI-T. This aligns with large-scale meta-analytic work demonstrating that anxiety disorders are reliably characterized by reductions in resting-state HRV, with effect sizes in the small-to-moderate range across panic disorder, generalized anxiety disorder, PTSD, and social anxiety disorder(79,135). RMSSD, often considered a vagal-specific index, trended toward a similar relationship with trait anxiety in our data, and importantly, both RMSSD and SDNN covaried within-person with fluctuations in state anxiety across sessions. These within-person effects suggest that short-term changes in vagal regulation may track day-to-day anxiety experience, consistent with the “autonomic inflexibility” models of anxiety(116). At the same time, baseline HRV did not predict anxiety ratings (APR, FPR) in response to our task nor moderate the effects of LIFU. This null result indicates that while HRV reflects a general vulnerability or endophenotypic marker of anxiety(80), it may not directly index acute responsiveness to neuromodulation in this context. Similar findings have been reported in experimental stress paradigms, where group-level HRV differences between anxious and non-anxious participants are robust at rest but less consistent during stressor exposure(136). Thus, HRV may be best conceptualized as a trait-like marker of anxiety risk and regulatory capacity rather than a dynamic predictor of task-evoked modulation. The distinction between RMSSD and SDNN is also worth noting. While RMSSD predominantly captures parasympathetic vagal tone, SDNN reflects overall variability across frequency bands and may better capture long-term autonomic balance(80,137). In our data, SDNN showed stronger cross-sectional correlations with both trait and state anxiety than RMSSD, suggesting it may be a more sensitive index of chronic anxiety severity.

Nevertheless, both indices supported the view that diminished HRV relates to heightened anxiety, in line with the neurovisceral integration and polyvagal models(116). Taken together, these findings reinforce the position that low HRV constitutes a physiological endophenotype of anxiety disorders(80). While many baseline factors have been demonstrated to affect non-invasive brain stimulation including physical activity, time of day and age (all factors that also affect HRV) (138) our results show that baseline HRV, while reflective of trait and state anxiety, does not moderate responsiveness to LIFU in the current design.

#### Baseline Effects: Heart Rate

Our results demonstrate that higher resting heart rate (HR) was associated with greater state and trait anxiety across participants, consistent with epidemiological and experimental findings linking elevated HR to anxiety and impaired vagal regulation. Both STAI-S and STAI-T showed significant positive associations with baseline HR, and ordinary least squares models indicated that a modest 5 bpm increase predicted roughly 1.5–1.7 points higher anxiety severity. These between-person associations were paralleled by within-person trends, whereby days with above-mean HR tended to coincide with elevated state anxiety, although these effects were weaker. Taken together, the data support the view that tonic increases in HR represent a physiological marker of anxiety severity. These results align with large cohort findings showing that generalized anxiety disorder is associated with small but robust elevations in resting HR alongside decreases in HRV(139). Importantly, increased resting HR has been identified as a correlate of diminished vagal activity and autonomic imbalance, which may help explain links between anxiety disorders and cardiovascular morbidity(139,140). Experimental studies similarly show that anxious individuals, particularly those with GAD and panic disorder, often exhibit elevated baseline HR and exaggerated HR responses to stressors such as hyperventilation(116), although findings vary depending on the disorder subtype and context. Our moderation models further revealed that baseline HR predicted anxiety-potentiated responding under LIFU, but only for modulation of the anterior insula (AI). Both between- and within-person HR components interacted with AI LIFU to predict APR ratings, whereas no such effects were observed for aMCC or sham conditions. This suggests that baseline cardiac state may confer sensitivity to insula-targeted modulation, consistent with the insula’s role in interoceptive monitoring of heartbeat and integration of autonomic signals with subjective anxiety (132,140). However, the absence of HR effects on FPR ratings indicates that this relationship may be specific to anticipatory processes rather than general fear responding. From a mechanistic perspective, elevated HR may index chronic sympathetic predominance and reduced parasympathetic flexibility, processes implicated in the pathophysiology of anxiety (116,140). While HR is less specific than HRV for vagal control, its predictive utility in our LIFU analyses highlights the potential importance of tonic cardiovascular state in shaping neuromodulatory outcomes. Clinically, this resonates with models suggesting that persistent high HR reflects not only a symptom of anxiety but also a risk factor for cardiovascular disease (139). These findings suggest that tonic cardiovascular activity may influence both subjective anxiety and responsiveness to targeted interventions. Future studies should further examine whether interventions that reduce resting HR may synergize with neuromodulation strategies to optimize outcomes.

### Summary and implications

Our findings demonstrate that low-intensity focused ultrasound can dissociate the neural substrates of anticipatory anxiety and cue-driven fear in healthy individuals with circuit-level precision. Modulation of the ventral anterior insula down-shifted arousal and strengthened brain–body– behavior coupling during uncertainty, while anterior mid-cingulate modulation selectively dampened cue-potentiated fear responses. These results align with predictive processing and salience network models in which the insula integrates interoceptive signals to shape anxious anticipation, and the aMCC mobilizes defensive actions to explicit threat. By causally linking these cortical hubs to distinct facets of threat reactivity, this work underscores the translational potential of ultrasound neuromodulation to parse and ultimately target anxiety phenotypes that differ in their reliance on uncertainty-driven versus cue-driven processes.

## Data Availability

All data produced in the present study are available upon reasonable request to the authors

