## Supplemental Data for "Anterior insula and mid-cingulate cortex differentially regulate anxiety and fear brain–body responses"

**Supplemental Table 1: Targets coordinates and depths**

|  |  | **MNI Coordinates** | | | | | | **Target Depth (mm)** | |
| --- | --- | --- | --- | --- | --- | --- | --- | --- | --- |
|  |  | **X** | **Y** | **Z** | **X** | **Y** | **Z** | **RAI** | **ACC** |
| **Male** | S1 | 35 | 14 | -14 | 0 | 22 | 26 | 41.9 | 50.5 |
|  | S2 | 33 | 37 | 6 | -1 | 26 | 37 | 41.4 | 50.5 |
|  | S3 | 37 | 19 | 0 | 0 | 22 | 26 | 37.6 | 49.2 |
|  | S4 | 33 | 28 | 1 | 0 | 22 | 26 | 36.6 | 49.1 |
|  | S5 | 38 | 9 | -10 | 0 | 22 | 26 | 41 | 54 |
|  | S6 | 32 | 20 | -2 | -5 | 18 | 36 | 43.6 | 44.1 |
|  | S7 | 37 | 21 | -1 | 0 | 22 | 26 | 36.9 | 49 |
|  | S8 | 37 | 18 | -1 | 0 | 22 | 26 | 41.7 | 49 |
|  | S9 | 34 | 22 | -8 | 0 | 22 | 26 | 37.9 | 50.9 |
|  | S10 | 38 | 15 | -11 | 0 | 22 | 26 | 35.7 | 56.6 |
|  | S11 | 31 | 27 | 6 | -6 | 21 | 18 | 38.1 | 49 |
|  | S12 | 40 | 10 | -11 | 0 | 22 | 26 | 35.3 | 46.7 |
| **Mean** |  |  |  |  |  |  |  | **39.0** | **49.9** |
| **SD** |  |  |  |  |  |  |  | **2.8** | **3.2** |
| **Female** | S13 | 36 | 14 | -12 | 0 | 22 | 26 | 35.2 | 49.9 |
|  | S14 | 33 | 21 | -4 | 0 | 20 | 26 | 44.2 | 45.2 |
|  | S15 | 40 | 5 | -10 | 0 | 22 | 26 | 33.8 | 44.9 |
|  | S16 | 32 | 14 | 6 | 0 | 22 | 26 | 43.4 | 49.4 |
|  | S17 | 35 | 11 | -13 | 0 | 22 | 26 | 37.9 | 46 |
|  | S18 | 37 | 13 | -13 | 0 | 22 | 26 | 37.2 | 51.8 |
|  | S19 | 41 | 10 | -12 | 0 | 22 | 26 | 37.4 | 43.9 |
|  | S20 | 36 | 9 | -12 | 0 | 22 | 26 | 38.7 | 46.8 |
|  | S21 | 35 | 18 | 2 | 0 | 22 | 26 | 40.7 | 46.8 |
|  | S22 | 36 | 12 | -11 | 0 | 22 | 26 | 31.3 | 44.9 |
|  | S23 | 36 | 14 | -8 | 0 | 22 | 26 | 37 | 45.1 |
|  | S24 | 40 | 11 | -13 | 0 | 22 | 26 | 33.7 | 47.1 |
|  | S25 | 32 | 21 | 7 | 0 | 22 | 26 | 39.9 | 44.6 |
|  | S26 | 38 | 11 | -9 | 0 | 22 | 26 | 31 | 46.1 |
|  | S27 | 37 | 12 | -11 | 0 | 22 | 26 | 34.9 | 45.7 |
|  | S28 | 35 | 28 | 4 | 0 | 22 | 26 | 35.8 | 50.7 |
|  | S29 | 33 | 26 | 12 | 0 | 26 | 28 | 33.1 | 40.1 |
|  | S30 | 39 | 9 | -14 | 0 | 22 | 26 | 40.5 | 46.6 |
|  | S31 | 40 | 6 | -11 | 0 | 22 | 26 | 33.1 | 48.6 |
|  | S32 | 35 | 20 | 0 | 0 | 22 | 26 | 36 | 48 |
|  | S33 | 41 | 11 | -12 | 0 | 22 | 26 | 34.2 | 45.5 |
|  | S34 | 35 | 16 | 10 | -2 | 29 | 22 | 39.2 | 45.6 |
|  | S35 | 38 | 7 | -14 | 0 | 22 | 26 | 36.9 | 46.3 |
|  | S36 | 36 | 19 | 10 | 0 | 22 | 26 | 32 | 47.5 |
|  | S37 | 39 | 10 | -9 | 0 | 22 | 26 | 33.6 | 48 |
|  | S38 | 41 | 10 | -8 | 0 | 22 | 26 | 37 | 50.7 |
|  | S39 | 39 | 11 | -11 | 0 | 22 | 26 | 33.2 | 43.5 |
|  | S40 | 31 | 20 | 7 | 0 | 22 | 26 | 36.9 | 47 |
| **Mean** |  |  |  |  |  |  |  | **36.4** | **46.7** |
| **SD** |  |  |  |  |  |  |  | **3.4** | **2.5** |

**Targeting Error**

**Supplementary Figure 1**

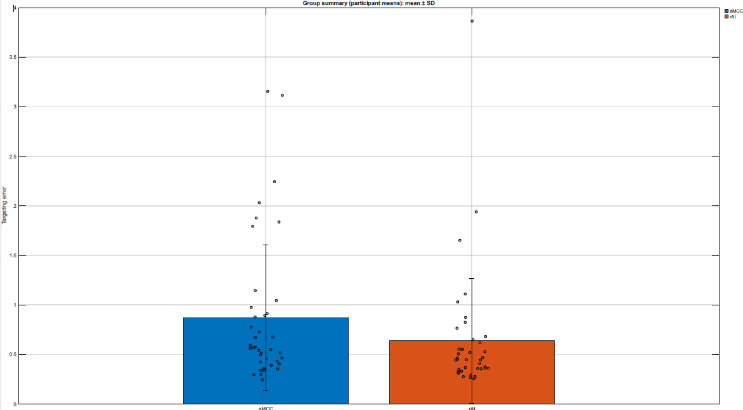

**Supplementary Figure 2**

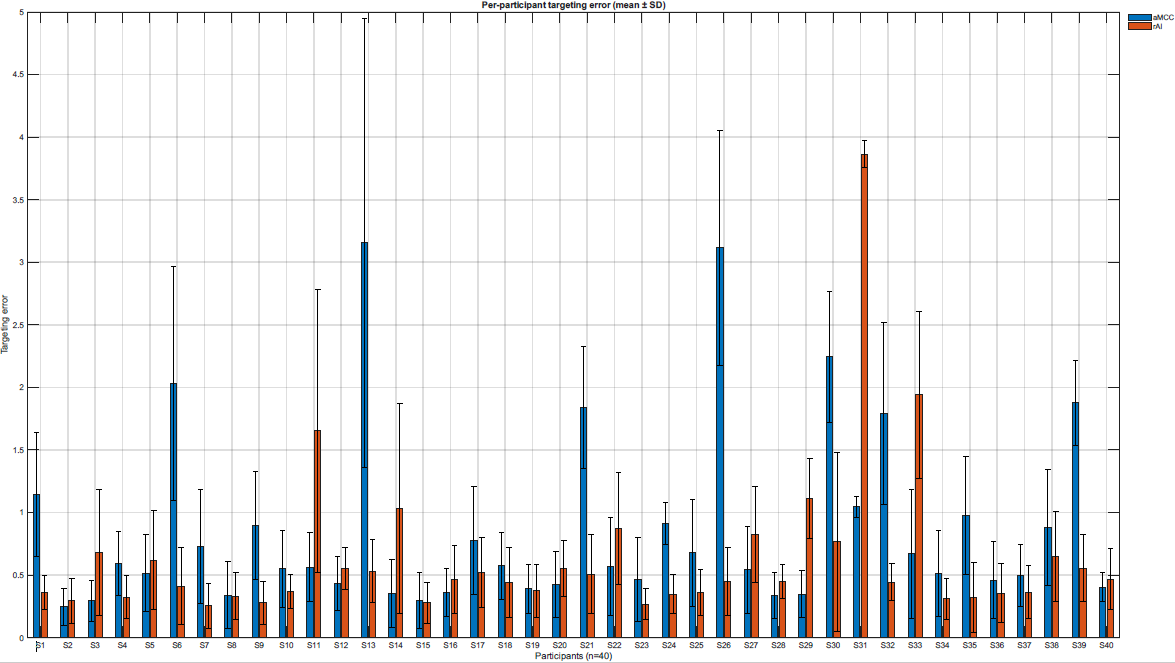

**Supplementary Figure 3**

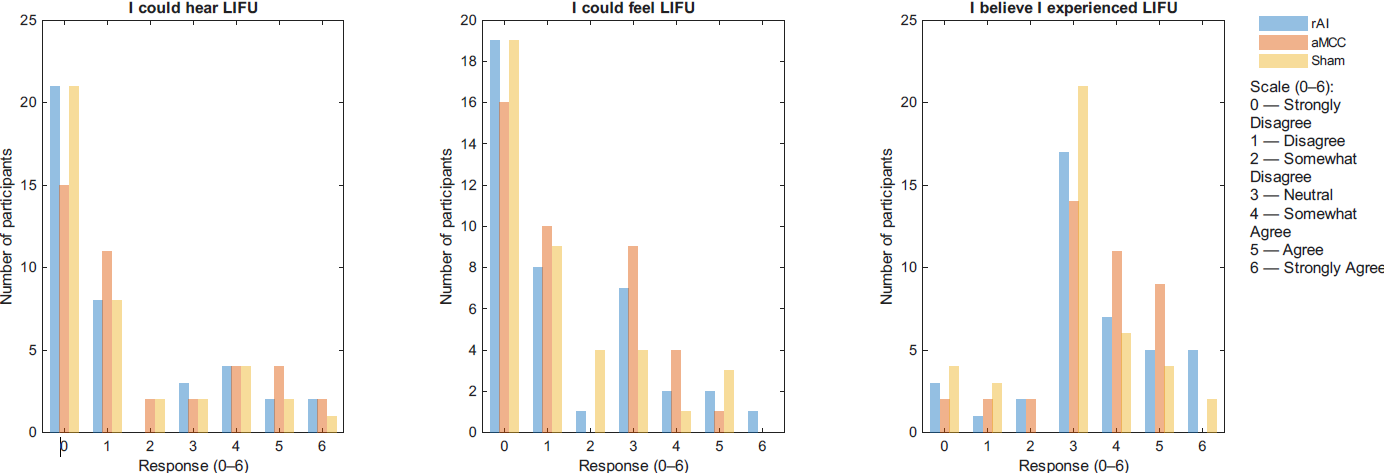

*Anxiety Ratings*

**Supplementary Table 2**. Raw anxiety ratings for NPU task for each LIFU condition. Data are mean ± SEM.

|  | **AI** | | **ACC** | | **Sham** | |
| --- | --- | --- | --- | --- | --- | --- |
|  | Cue | No Cue | Cue | No Cue | Cue | No Cue |
| **Neutral** | 1.1 ± 0.2 | 0.8 ± 0.2 | 1.1 ± 0.2 | 0.7 ± 0.1 | 1.3 ± 0.2 | 0.9 ± 0.2 |
| **Predictable** | 4.0 ± 0.4 | 2.1 ± 0.3 | 3.6 ± 0.4 | 1.9 ± 0.3 | 4.7 ± 0.5 | 2.0 ± 0.3 |
| **Unpredictable** | 4.1 ± 0.4 | 2.4 ± 0.3 | 3.8 ± 0.4 | 3.4 ± 0.3 | 4.1 ± 0.4 | 3.7 ± 0.4 |

*EMG Startle Response***.**

**Supplementary Table 3**. Raw EMG AUC for NPU task for each LIFU condition. Data are mean ± SEM.

|  | **AI** | | **ACC** | | **Sham** | |
| --- | --- | --- | --- | --- | --- | --- |
|  | Cue | No Cue | Cue | No Cue | Cue | No Cue |
| **Neutral** | 73.1 ± 6.1 | 72.6 ± 6.9 | 69.3 ± 7.0 | 72.4 ± 7.5 | 73.2 ± 6.3 | 73.8 ± 6.4 |
| **Predictable** | 120.8 ± 10.5 | 101.4 ± 8.8 | 105.3 ± 11.8 | 92.9 ± 9.5 | 115.3 ± 10.9 | 95.2 ± 9.3 |
| **Unpredictable** | 102.2 ± 8.7 | 77.1 ± 10.1 | 93.4 ± 9.9 | 99.4 ± 9.2 | 100.5 ± 10.7 | 94.6 ± 8.1 |

**Supplementary Figure 4**

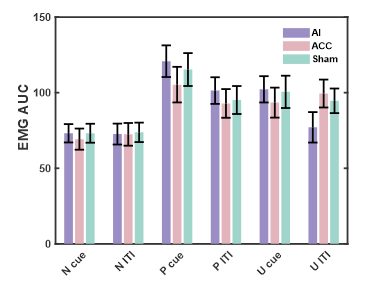

*Electrodermal Response.*

**Supplementary Table 4**. EDR AUC for NPU task for each LIFU condition. Data are mean ± SEM.

|  | **AI** | | **ACC** | | **Sham** | |
| --- | --- | --- | --- | --- | --- | --- |
|  | Cue | No Cue | Cue | No Cue | Cue | No Cue |
| **Neutral** | 0.006 ± 0.001 | 0.007 ± 0.002 | 0.004 ± 0.001 | 0.005 ± 0.001 | 0.009 ± 0.003 | 0.006 ± 0.002 |
| **Predictable** | 0.016 ± 0.003 | 0.012 ± 0.002 | 0.013 ± 0.002 | 0.008 ± 0.001 | 0.013 ± 0.003 | 0.009 ± 0.002 |
| **Unpredictable** | 0.016 ± 0.003 | 0.013 ± 0.003 | 0.013 ± 0.002 | 0.015 ± 0.004 | 0.012 ± 0.002 | 0.013 ± 0.004 |

**Supplementary Figure 5**

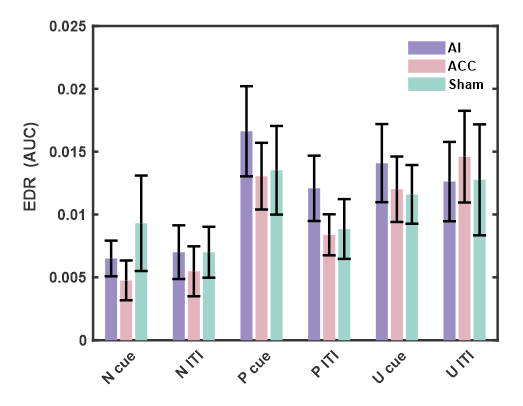

We fit linear mixed-effects models with fixed effects of LIFU (AI, ACC, Sham), TASK (APR, FPR) and their interaction, and random intercepts and TASK slopes by subject: EDR ~ LIFU * TASK + (1 + TASK | Subject).

We fit linear mixed-effects models with fixed effects of LIFU (AI, ACC, Sham), TASK (APR, FPR), and their interaction, and random intercepts and TASK slopes by subject: EDR ~ LIFU × TASK + (1 + TASK | Subject).

The model showed a significant intercept (F(1,87.4) = 5.75, p = 0.019), indicating that average EDR responses were greater than zero, but there were no significant main effects of LIFU (F(2,200) = 1.33, p = 0.267), TASK (F(1,145.4) = 0.16, p = 0.687), or their interaction (F(2,200) = 0.64, p = 0.528).

Planned contrasts confirmed the absence of LIFU effects. During APR, neither AI (est = –0.14, SE = 2.44, t(234) = –0.06, p = 0.953, BH-q = 0.953) nor ACC stimulation (est = 3.36, SE = 2.44, t(234) = 1.38, p = 0.169, BH-q = 0.169) differed significantly from Sham. Similarly, during FPR, both AI (est = –0.13, p = 0.956) and ACC (est = –0.01, p = 0.998) did not differ from Sham.

Estimated marginal means illustrated this null pattern: Sham responses were comparable across APR (M = 5.76, 95% CI [1.03, 10.49]) and FPR (M = 4.67, 95% CI [1.13, 8.22]). AI stimulation produced nearly identical responses (APR M = 5.62; FPR M = 4.54), while ACC stimulation numerically increased APR responses (M = 9.12, 95% CI [4.39, 13.86]) but not FPR (M = 4.67, 95% CI [1.12, 8.22]), though these differences were not statistically reliable.

Together, these results indicate that EDR responses were not significantly modulated by LIFU or task condition, in contrast to the EMG findings.

*Heart Rate.*

**Supplementary Table 5**. Heart Rate for NPU task for each LIFU condition. Data are mean ± SEM in beats per minute (BPM)

|  | **AI** | | **ACC** | | **Sham** | |
| --- | --- | --- | --- | --- | --- | --- |
|  | Cue | No Cue | Cue | No Cue | Cue | No Cue |
| **Neutral** | 70.9 ± 2.3 | 71.3 ± 2.2 | 70.8 ± 1.7 | 71.4 ± 1.7 | 71.4 ± 1.6 | 71.8 ± 1.6 |
| **Predictable** | 70.9 ± 2.2 | 72.4 ± 2.3 | 70.0 ± 1.7 | 71.7 ± 1.7 | 71.8 ± 1.7 | 72.0 ± 1.6 |
| **Unpredictable** | 70.8 ± 2.2 | 70.0 ± 2.2 | 70.4 ± 1.7 | 71.3 ± 1.8 | 70.5 ± 1.6 | 71.5 ± 1.7 |

**Supplementary Figure 6**

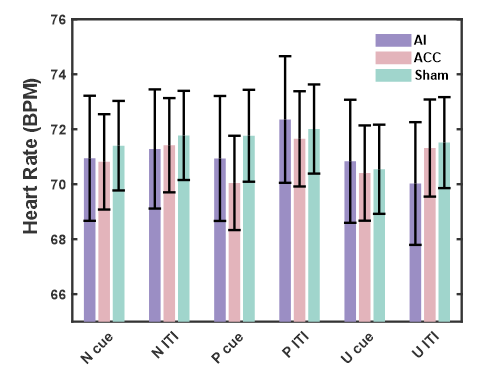

*EEG. Startle.*

**Supplementary Table 6**. Peak-to-peak ERP amplitude for NPU task for each LIFU condition. Data are mean ± SEM in microvolts (µV)

|  | **AI** | | **ACC** | | **Sham** | |
| --- | --- | --- | --- | --- | --- | --- |
|  | Cue | No Cue | Cue | No Cue | Cue | No Cue |
| **Neutral** | 18.0 ± 1.7 | 16.8 ± 1.3 | 17.4 ± 1.1 | 16.6 ± 1.0 | 17.6 ± 1.0 | 16.8 ± 0.9 |
| **Predictable** | 17.3 ± 1.3 | 18.9 ± 1.7 | 17.8 ± 1.3 | 18.8 ± 1.2 | 20.4 ± 1.2 | 18.2 ± 1.0 |
| **Unpredictable** | 19.2 ± 1.6 | 17.1 ± 1.4 | 18.9 ± 1.1 | 18.6 ± 1.3 | 18.8 ± 1.0 | 17.8 ± 0.9 |

**Supplementary Figure 7**

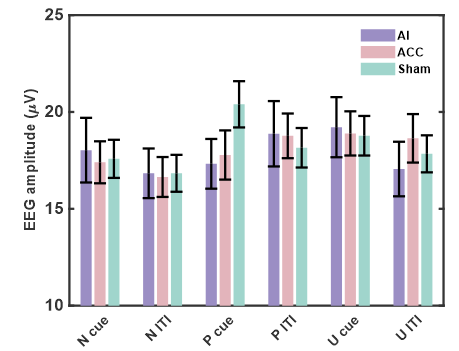

**Brain-Body-Behavior Coupling**

*EMG startle*

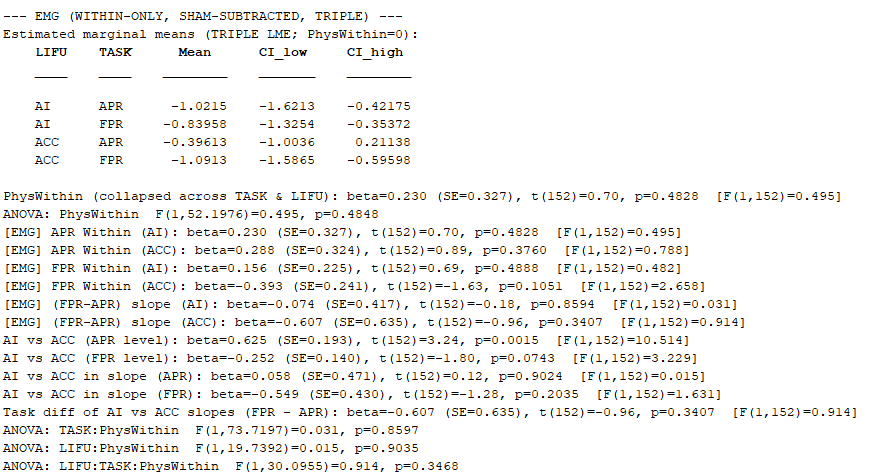

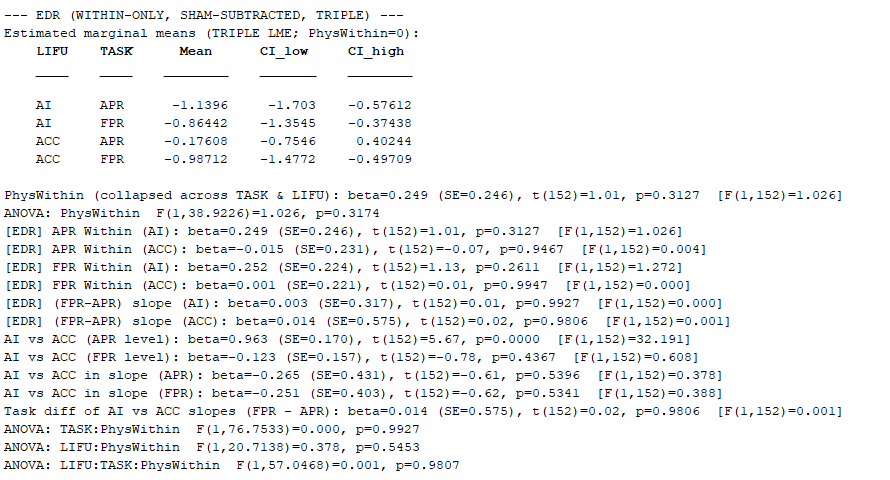
*Electrodermal Response (EDR)*

Heart Rate

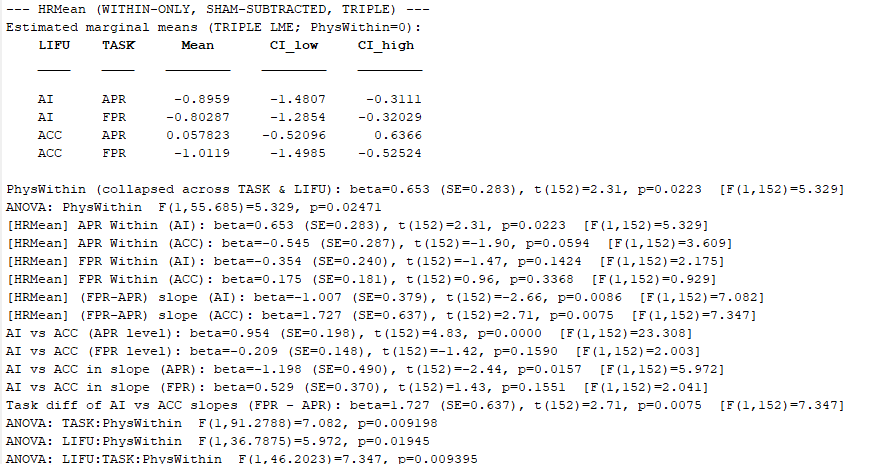

EEG

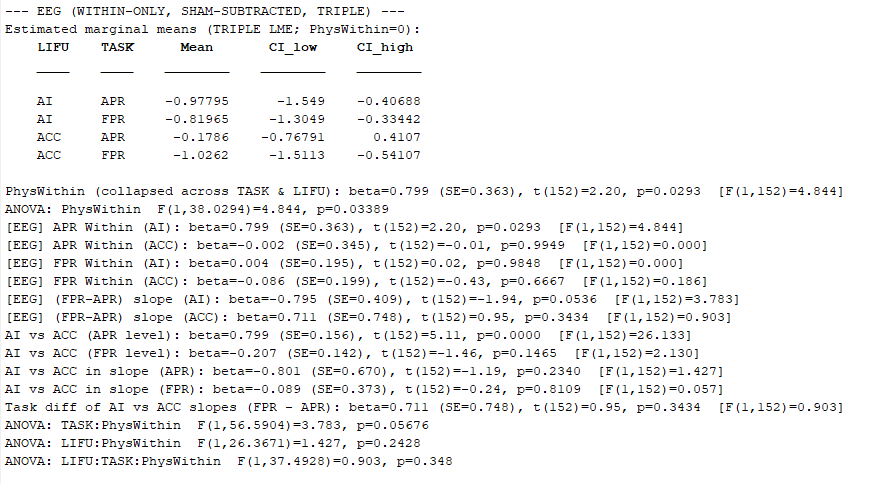
